# Disruption of CTCF binding by germline non-coding variants in *CDKN2B* suppress *CDKN2A* expression and predispose to melanoma

**DOI:** 10.64898/2026.06.01.26352322

**Authors:** Jessica L Scales, Jayne A Barbour, Alisa M Goldstein, Rebecca Hennessey, Mai Xu, Abigail J Dennis, Sophie Papiernik, Jung Kim, Sudipto Das, Haocheng Yang, S. Chul Kwon, Kornelia Gladysz, Rohit Thakur, Joshuah Yon, Linh Bui-Raborn, Douglas R Stewart, Raj Chari, Paula L Hyland, Jiyeon Choi, Tongwu Zhang, Wen Luo, Kedest Teferi, Thorkell Andresson, Xin Li, Kristine M Jones, Amy Hutchinson, Belynda D Hicks, W. Ryan Diver, Adriana Lori, Steven C Moore, Margaret A Tucker, Michael R Sargen, Kevin M Brown, Jason W H Wong, Xiaohong R Yang

**Affiliations:** Division of Cancer Epidemiology and Genetics, National Cancer Institute, Rockville, MD, USA; School of Biomedical Sciences, The University of Hong Kong, Hong Kong SAR, China; Oncology and Immunology, Hong Kong Science Park, Hong Kong SAR, China; CCR Protein and Metabolite Characterization Core (PMCC), Frederick National Laboratory for Cancer Research, National Cancer Institute, Frederick, MD, USA; Genome Modification Core, Laboratory Animal Sciences Program, Frederick National Laboratory for Cancer Research, Frederick, MD, USA; Division of Applied Regulatory Science, Center for Drug Evaluation and Research, US Food and Drug Administration, Chevy Chase, MD, USA; Cancer Genomics Research Laboratory, Frederick National Laboratory for Cancer Research, Frederick, MD, USA; Department of Population Science, American Cancer Society, Atlanta, GA, USA

## Abstract

Some melanoma-prone families linked to the 9p21 locus, harboring the established susceptibility gene *CDKN2A*, lack pathogenic protein-coding variants. Using whole-exome and targeted sequencing, we identified three rare single-nucleotide variants in two melanoma-prone families and one sporadic melanoma case. Variants map to a conserved CTCF-bound region within the first intron of *CDKN2B* that physically interacts with *CDKN2A*. Analysis of UK Biobank showed significant enrichment of variants in this region in melanoma cases. Variants result in diminished CTCF binding *in vitro*. CTCF ChIP-seq in fibroblasts from the carriers of the largest family demonstrated loss of CTCF binding, accompanied by weakened promoter interactions and allele-specific reduction of *CDKN2A* p16 transcript expression from the variant haplotype. CRISPR-based perturbation of this region and editing of the large family variant into melanocytes resulted in reduced expression of p14 and p16 *CDKN2A* transcripts. These findings suggest that non-coding regulatory variants function as high-penetrance susceptibility alleles in melanoma families by altering *CDKN2A* function.

## Introduction

Familial cancers represent approximately 5-10% of all cancers and are attributed to high-penetrance germline variants that predispose to disease [1]. Consistent with this, cutaneous melanoma is highly heritable [2, 3]. Roughly 10% of cases occur in a familial setting where ∼40% of families are explained by pathogenic germline variants in the *CDKN2A* gene on chromosome band 9p21 [4–11]. A further ∼10% of families are explained by rarer pathogenic variants in a handful of other genes elsewhere in the genome, including *CDK4* [12–16], the promoter of *TERT* [17, 18], and shelterin complex members *POT1* [19, 20], *ACD* [21], *TERF2IP* [21], and *TINF2* [22].

Of the remaining families, a sizable proportion are nonetheless linked to 9p21 [23, 24], including families with early-onset cases, members with multiple primary melanomas, large numbers of melanocytic nevi, and dysplastic nevus diagnoses, which are hallmarks of families carrying germline *CDKN2A* pathogenic variants (PVs) [7, 9, 25]. This has led to the hypothesis that such families may be explained by non-coding germline variation resulting in inactivation of *CDKN2A*. While most common cancer-predisposing variants identified via genome-wide association studies (GWAS) are thought to be non-coding [26], there are few examples of high-penetrance non-coding variants occurring in the context of multi-case cancer families, with rare germline promoter mutations in *TERT* found in melanoma families being a notable example [17, 18]. To date, relatively few non-coding familial risk variants around *CDKN2A* have been identified, with only a rare intronic *CDKN2A* variant [27] and some large germline copy number changes directly over *CDKN2A* [28, 29] reported.

Alternatively, many have speculated that these families could be explained by variants in a second susceptibility gene located at 9p21. Studies of DNA copy number and loss of heterozygosity (LOH) at 9p21 in melanoma tumors are consistent with this hypothesis, with LOH in tumors frequently extending well beyond *CDKN2A* and overlapping numerous other genes in the region [30–36], including clusters that do not overlap *CDKN2A* itself [37]. Among the genes in the region, *CDKN2B* represents perhaps the strongest *a priori* candidate, encoding p15 (Ink4b), a family member of the p16 protein encoded by *CDKN2A* (Ink4a), which is an inhibitor of cyclin dependent kinases 4 and 6 (CDK4/CDK6), and an established tumor suppressor gene [24, 38, 39]. In melanocytes, loss of *CDKN2B* has been shown to promote progression from benign nevus to tumor [40] and may be involved in centrosome overduplication [41]. Despite being an attractive candidate familial melanoma gene, numerous targeted [24, 42–45] as well as genome- or exome-wide studies of high-risk melanoma families [19, 20, 46–48] or sporadic cases [49] have failed to establish *CDKN2B* as a susceptibility gene to date. The possibility of variants proximal to *CDKN2B* regulating *CDKN2A* via long-range interactions has not been explored.

Here, we follow up previous whole-exome sequencing (WES) findings from high-risk melanoma families from the United States [46] where we identified two families harboring distinct nearby rare variants within a single consensus CTCF-binding motif, which also alter the protein-coding sequence of a non-canonical transcript of *CDKN2B* (p10). We identified a third such variant in WES data from a melanoma case-control study [47]. We show that these variants are unlikely to function via alteration of *CDKN2B* protein-coding sequence, as the isoform is not expressed in melanocytes or melanoma. Instead, we experimentally show reduced CTCF binding resulting from one of these variants coupled with allele-specific silencing of *CDKN2A* expression, suggesting that these variants function via altered *cis*-regulation of *CDKN2A*. Finally, we independently validated this finding via CRISPR inhibition and editing. From UK Biobank, we examined 469,376 individuals to corroborate that the variant confers high-risk for melanoma, providing an example of a germline cancer variant that cannot be explained through the lens of protein-coding biology.

## Results

### Identification of rare, cosegregating CDKN2B variants in high-risk melanoma families

We previously reported a large melanoma-prone family with characteristics consistent with those harboring *CDKN2A* germline pathogenic variants (current status: 14 members with melanoma, median age at melanoma diagnosis = 39 years, 3 members with multiple primary melanoma [MPM]) (pedigree M, **Figure 1**) and evidence of linkage to 9p21 [5]. However, no *CDKN2A* pathogenic variant was identified even after ensuing evaluation of copy number variants, large deletions, or exon 1b [28, 46, 50]. Recent long-read whole genome sequencing of two 9p21-linked melanoma patients confirmed the absence of structural variants in the 9p21 region (data not shown). Subsequent examination using whole exome sequencing (WES) uncovered a rare protein-coding change to a non-canonical transcript of *CDKN2B*: (chr9 g.22008768C>G, hg38, RefSeq NM_078487 p.Ala64Thr; gnomAD 4.1.0 non-Finnish European/NFE allele frequency = 5.09 x 10^-6^) [46]. A second smaller family (pedigree X: 3 melanoma cases, median age at melanoma diagnosis – 43 years; 1 member with MPM; **Figure 1**) also harbored a similar type of *CDKN2B* variant (chr9 g.22008764C>T, RefSeq NM_078487 p.Gln62His, gnomAD NFE AF = 2.12 x 10^-6^) [46]. These variants were technically validated by Sanger sequencing [47]. In addition, assessment of gene panel sequencing data [47] from sporadic melanoma cases within the Prostate, Lung, Colon, Ovary (PLCO) screening trial identified a case harboring a third such variant nearby (chr9 g.22008773G>C, NM_078487 p.Arg61Gly, gnomAD NFE AF = 8.49 x 10^-7^).

**Figure 1.**
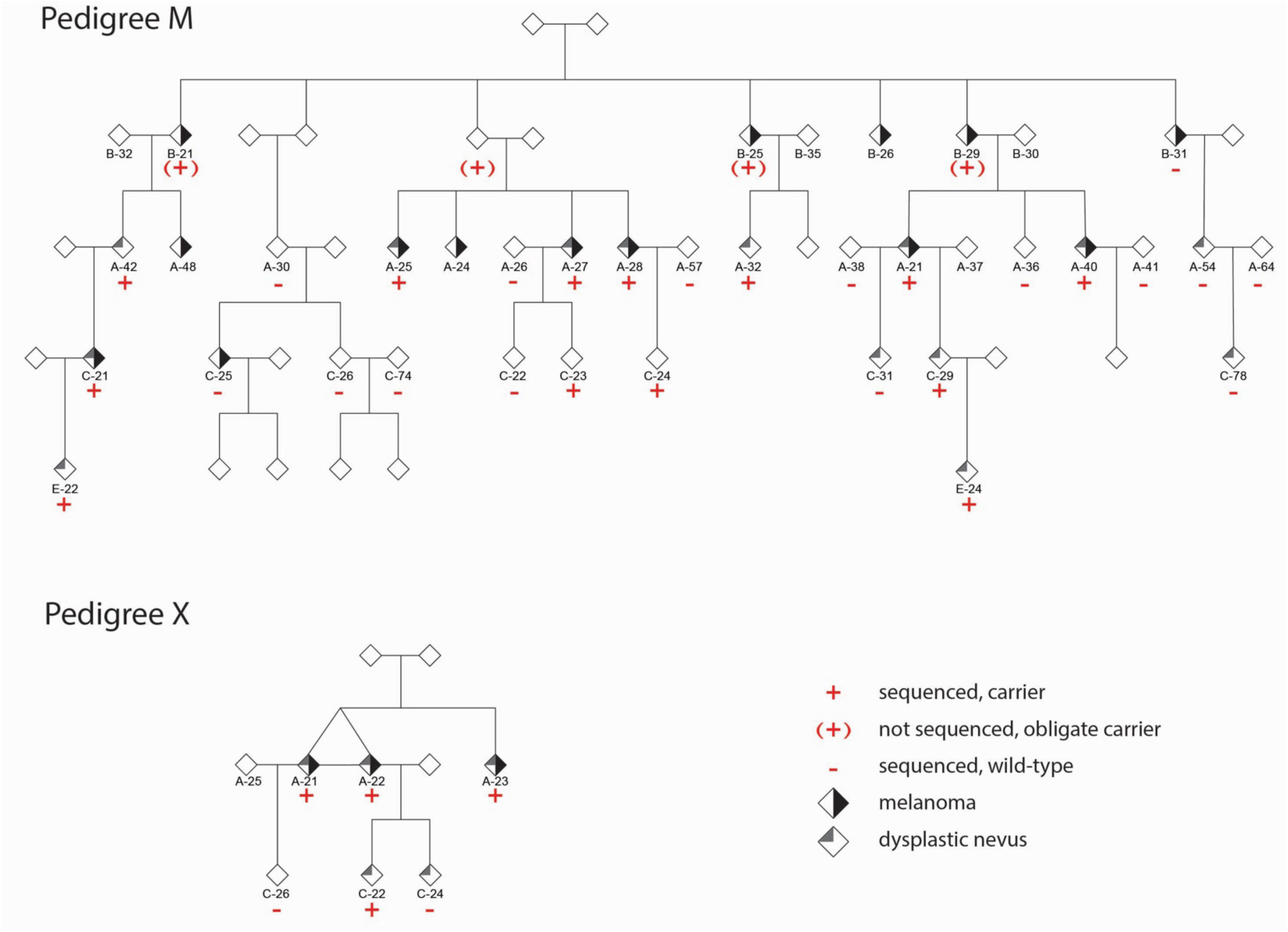
Pedigrees carrying *CDKN2B* variants. Directly sequenced variant carriers are denoted by “+”, sequenced non-carriers by “-“, and obligate carriers by “(+)”. Melanoma case status, as well as dysplastic nevus diagnoses are denoted (see legend).

Gene panel sequencing of family members with available DNA from both pedigrees M and X showed evidence of co-segregation of each respective *CDKN2B* variant with melanoma with incomplete penetrance (**Figure 1**), a pattern that is consistent with families with *CDKN2A* pathogenic variants. All three melanoma cases in pedigree X harbored g.22008764C>T, while 9 of 11 cases of the larger 14-case pedigree M harbored or were obligate carriers of the g.22008768C>G variant (**Figure 1**). Among the two individuals (B-31 and C-25) not carrying the variant (i.e. phenocopies), B-31 had melanoma-*in-situ* diagnosed in the 80s. C-25 also had a later age at diagnosis (50-55 years) than the melanoma cases harboring the variant (median age at diagnosis for cases with known variant status: 38 years).

Members of both families tended to have high nevus counts, dysplastic nevi, and among melanoma cases, both an early age at diagnosis and MPM (**Supplementary Table 1**). For pedigree M, amongst directly-sequenced *CDKN2B* variant carriers with phenotyping data (n=13, 6 melanoma cases), all 6 melanoma cases had high numbers of melanocytic nevi and dysplastic nevi. Of the non-melanoma variant carriers, 2 had a “high”, 2 “medium”, and 3 “low” nevus count and the majority had dysplastic nevi (5/7). In contrast, the two melanoma cases without the *CDKN2B* variant had neither dysplastic nevi or high nevus counts. In the smaller family (pedigree X), the four identified carriers had a high nevus count and dysplastic nevi (**Supplementary Table 1** and **Figure 1**).

### Protein-coding function does not explain the risk-associated CDKN2B variants

All three germline *CDKN2B* variants are located within 10 bp of each other, and while all are annotated as protein-coding for the non-canonical p10 *CDKN2B* transcript (RefSeq NM_078487) [51–53], both are intronic for the canonical p15 (Ink4b) transcript (RefSeq NM_004936) with well-established tumor suppressor function including in melanoma (**Figure 2A**). We assessed potential pathogenicity of these variants on p10 protein function using a suite of prediction tools (**Supplementary Table 2**). While AlphaMissense [54] predicted all three variants as benign, roughly half of the algorithms predicted each of the variants as potentially pathogenic (8/15, 7/15, and 9/15 for g.22008764C>T, g.22008768C>G, and g.22008773G>C, respectively), suggesting potential impact on the function of p10. Thus, we also assessed the relative expression of *CDKN2B* isoforms from RNA sequencing (RNA-seq) data from melanocytic cells, including human primary melanocytes (n=106) [55], melanoma cell lines (n=75) [56], and melanoma tumors (n=445) [30] by assessing junction reads supporting each transcript. Junction reads supporting p15 expression represented the vast majority of reads in all three data sets, respectively (99.5%, 98.7%, and 99.2%; **Figure 2B-C; Supplementary Table 3**). In primary melanocytes, only 3% of samples had a read supporting p10 expression (5 reads total); this number was slightly higher in melanoma cell lines and tumors (20 reads collectively in 22% of cell lines; 78 reads collectively in 13% of tumors). Finally, we assessed isoform expression in immortalized human melanocytes (C283Ts) via quantitative RT-PCR and observed little to no expression of the p10/NM_078487 isoform with fold-difference of p15 versus p10 ranging from 103-to 207-fold across four biological replicates (**Supplementary Figure 1**). These data suggest that while the three variants may conceivably alter p10 function, very little of this *CDKN2B* transcript is expressed in melanocytic cells.

**Figure 2.**
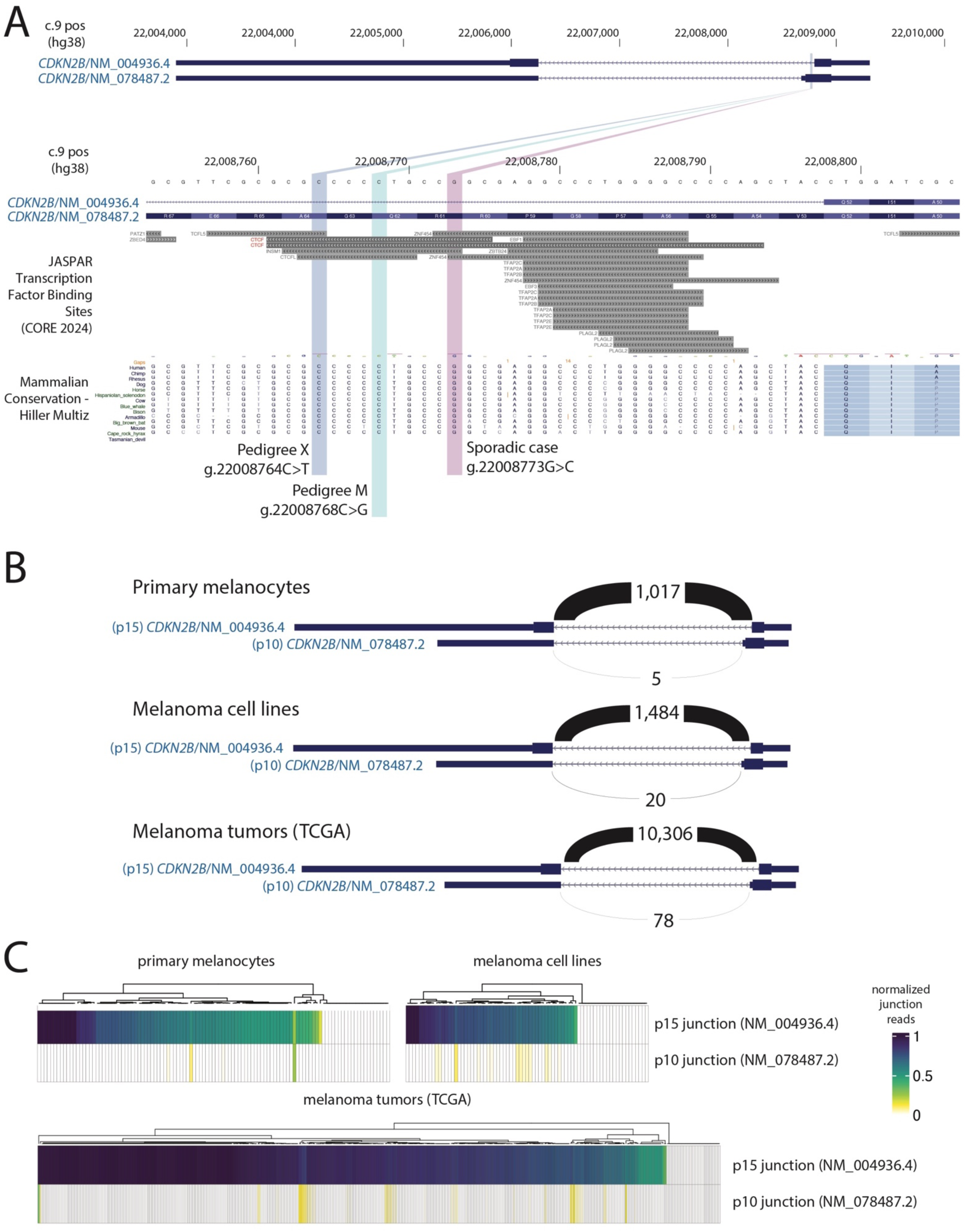
(**A**) Genomic location of risk-associated *CDKN2B* variants. The locations of three variants found in pedigrees M, X, and a sporadic melanoma case are shown relative to the canonical p15 (NM_004936.4) and alternative p10 transcripts (NM_078487.2) of *CDKN2B*. Transcription factor binding sites from the JASPAR Core 2024 collection (including for CTCF) are shown, as is conservation of bases in the region across mammalian species. Coordinates are hg38. (**B**) Analysis of RNA sequencing junction reads from human primary melanocytes (n=106), human melanoma cell lines (n=75), and human melanoma tumors (n=445) for the p15 and p10 transcripts of *CDKN2B*. Total number of junction reads supporting each transcript are shown. (**C**) Visualization of normalized junction read usage for individual melanocyte cultures, melanoma cell lines, and melanoma tumors; junction reads were normalized to the total read count 3’ of the splice acceptor site common to both transcripts (relative to the *CDKN2B* transcript).

### Risk-associated variants are located within a consensus CTCF-binding motif

Given that the variants are unlikely to operate through a protein-coding mechanism, we hypothesized that they may play a *cis*-regulatory role. To this end, we leveraged the deep learning model AlphaGenome [57], which makes regulatory variant effect predictions. AlphaGenome was run for the reference and mutant alleles for all three variants over a 1 Mb window, modelling regulatory effects based on dermal fibroblast data for transcription (RNA-sequencing), chromatin accessibility (DNAse I hypersensitivity sequencing; DHS), CTCF binding (CTCF Chromatin Immunoprecipitation sequencing; ChIP-seq), and active enhancers (H3K27Ac ChIP-seq). While little difference was apparent in the RNA-seq and H3K27Ac tracks, a striking difference was apparent in CTCF ChIP-seq predictions at the site of all three variants between variant and reference alleles (**Figure 3A-B**, **Supplementary Figures 2 and 3**), suggesting a substantial impact on CTCF binding. The model also revealed a loss of a small chromatin accessibility peak (DHS) specifically at the site of the variants without disrupting the larger DHS peak immediately downstream of the variant or elsewhere in the locus (**Figure 3A, Supplementary Figures 2 and 3**).

**Figure 3.**
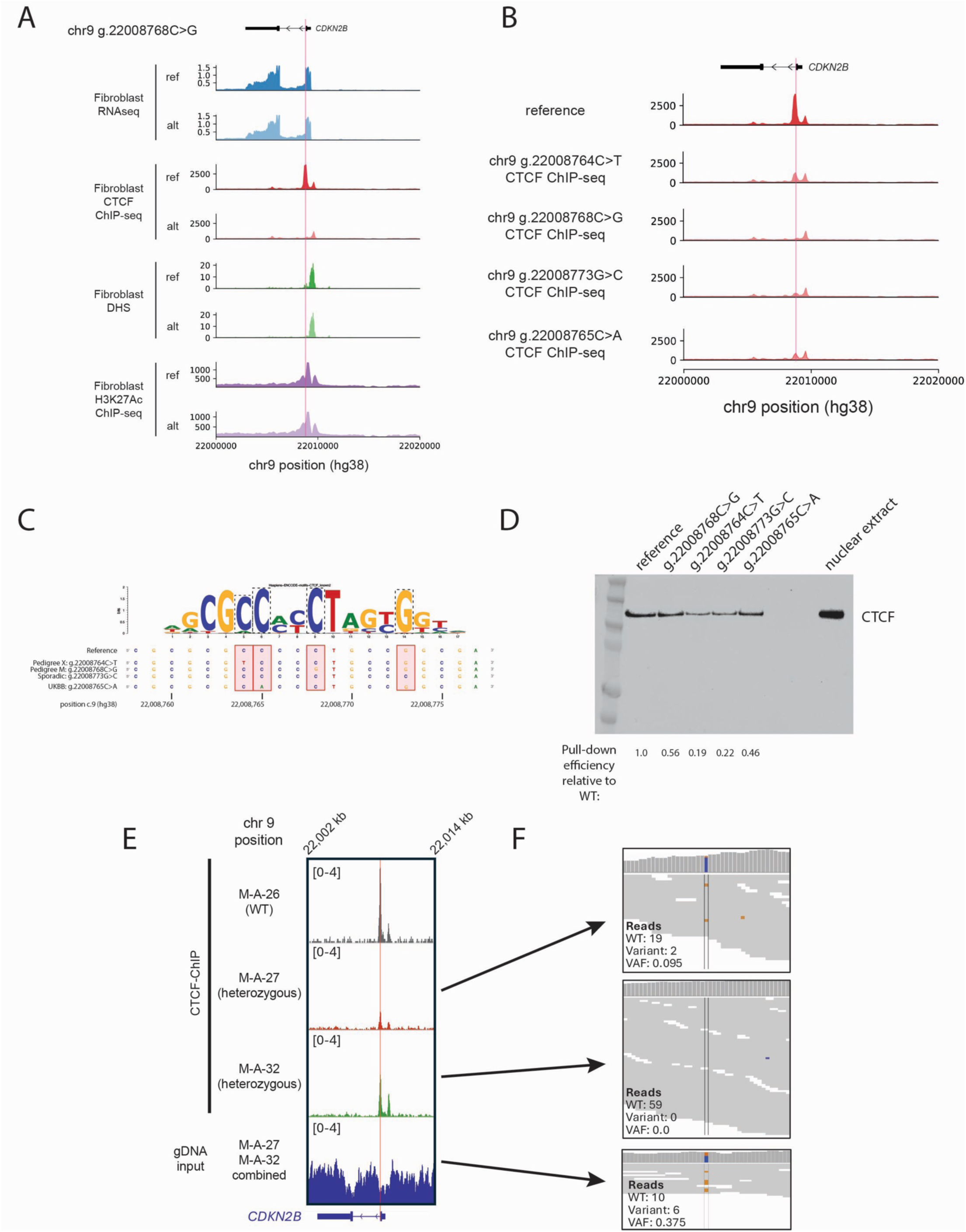
*CDKN2B* variants are located within a conserved CTCF binding site and alter CTCF binding. (A) AlphaGenome predictions of the consequences of the variant carried by pedigree M (chr9 g.22008768C>G) on transcription (RNA-seq), CTCF-binding (CTCF ChIP-seq), chromatin accessibility as assessed by DNAseI hypersensitivity sequencing (DHS), and histone mark H3K27Ac (H3K27Ac ChIP-seq). The predictions suggest predicted loss of CTCF binding as well as a modest reduction of chromatin accessibility over the variant position for the alternative allele (alt) relative to the reference (ref). Variant position is highlighted in pink. (**B**) AlphaGenome predictions of CTCF DNA-binding for all variants discovered in American melanoma cases and UKBB for reference and alternative alleles; variant position is highlighted in pink. (**C**) The location of the three *CDKN2B* variants are shown relative to a CTCF binding motif. Letter height reflects relative conservation of a given base within CTCF binding sites (generated by motifbreakR). Coordinates are based on hg38. (**D**) CTCF western after pulldown by oligonucleotides carrying non-coding variants. An oligonucleotide pulldown assay was performed using nuclear extract from immortalized C283T melanocytes using oligos carrying reference or variant sequences. Equal amounts of supernatant were loaded onto the protein gel following pulldown, as well as the nuclear extract used for the pulldown assays. The intensity of each band was determined by Image Lab software and pulldown efficiency was calculated by normalizing the band intensity with that of reference sequence oligo. Figure is representative of three replicate experiments. (**E**) CTCF ChIP-seq (and input) coverage over *CDKN2B* in fibroblasts from two variant carriers from pedigree M (M-A-27, M-A-32), as well as an unaffected married-in non-carrier (M-A-26). Pink shading reflects the location of the chr9 g.22008768C>G variant. Coordinates are hg38. Coverage is normalized using reads per genomic content (RPGC). (**F**) Variant and reference (WT) reads over chr9 g.22008768 in CTCF ChIP-seq in the heterozygous carriers (upper panels) and input (lower panel; whole-genome sequencing; WGS) libraries. Input WGS reads for affected carriers M-A-27 and M-A-32 combined are shown. Variant allele frequency (VAF) of CTCF ChIP (M-A-27 and M-A-32 combined) is significantly lower than the input (VAF: 0.025 versus 0.375, *P* = 0.0002, Fisher’s exact test) indicating allele specific loss of CTCF binding.

Consistent with these data, all three variants are located within highly conserved positions of a single predicted CTCF-bound region in the first intron of p15 (**Figure 3C**). Assessment of potential allele-specific impacts of all three variants on predicted binding of sequence-specific DNA binding proteins using motifbreakR (**Supplementary Table 4**) [58, 59] revealed that amongst the motifs most consistently interrupted by the variant alleles were CTCF-binding motifs (**Figure 3C**), with most showing predicted loss of binding for all three variants. Structural analyses of the CTCF-DNA complex previously reported that the positions in the motif corresponding to the variants are critical for CTCF-DNA binding [60]. For example, R395 of CTCF (protein database ID 5KKQ) forms hydrogen bonds with guanine position 8 on the complementary strand of the motif. This interaction becomes abrogated when the guanine at this position is replaced by cytosine with the chr9 g.22008768C>G variant found in pedigree M (**Supplementary Figure 4**). Likewise, one of the two hydrogen bonds from K364 formed with the guanine position 12 is lost when replaced with adenosine (pedigree X, g.22008764C>T), and the single hydrogen bond between D450 and the cytosine at position 3 is lost following substitution with guanine (sporadic case from PLCO, g.22008773G>C; **Supplementary Figure 4**).

### Association of CDKN2B CTCF-motif variants with melanoma risk

Considering these data suggest multiple risk-associated variants located within a single CTCF binding motif, we hypothesized that more broadly, genetic variation within the CTCF motif harboring these variants may be associated with melanoma risk. Specifically, we investigated a potential association in the UK Biobank (UKBB), comparing cancer-free controls to melanoma cases, restricting the analysis to only those variants overlapping the CTCF motif (chr9:22008761-22008775; hg38). While the number of carriers is small, we observe a significant enrichment of CTCF-motif variants in melanoma cases (0.036% in cases vs. 0.006% in controls, Fisher’s exact test *P* = 0.012; **Table 1**). This association was highly significant after adjusting for age and sex (*P* = 8.56 x 10^-4^; adjusted odds ratio [OR] = 8.34, 95% confidence interval [CI] = 2.40-29.04). The association with melanoma was likewise significant when restricting the analysis to the most commonly observed variant discovered in pedigree M (g.22008768C>G; Fisher’s *P* = 0.021; adjusted *P* = 1.79 x 10^-3^; adjusted OR = 12.66, 95% CI = 2.57-62.31) or any one of the three discovery variants (Fisher’s *P* = 0.031; adjusted *P* = 5.21 x 10^-3^; adjusted OR = 9.10, 95% CI = 1.93-42.87). Notably, the observed effect sizes are consistent with those expected for pathogenic variants in high-penetrance cancer susceptibility genes, including those previously reported for pathogenic *BRCA1* and *BRCA2* variants on breast cancer risk in UKBB (*BRCA1* OR = 8.73, 95% CI = 7.47–10.20; *BRCA2* OR = 5.68, 95% CI = 5.13–6.30) [61], as well as for pathogenic variants in *CDKN2A* on melanoma risk (OR = 12.8, 95% CI = 8.9-18.3) [62]. Of note, of the three melanoma cases from UKBB carrying a variant anywhere in the motif (mean age 66.7 years), two also had diagnosed nevi of the trunk and eyelid, respectively (ICD10 D22.5 and D22.1), consistent with the nevus phenotype described in the pedigrees. Two of the cases carried g.22008768C>G (pedigree M variant), while notably one carried a different rare variant, chr9 g.22008765C>A (gnomAD NFE AF = 1.70 x 10^-6^), which unlike the variants described previously, is synonymous for the p10 *CDKN2B* transcript (and intronic for p15). This variant is located within one of the most highly conserved positions in the CTCF motif (**Figure 3C**), with predicted loss of CTCF binding (**Supplementary Table 4**) via disruption of a hydrogen bond with R367 (**Supplementary Figure 4**), as well as strong effects on predicted CTCF-binding by AlphaGenome (**Figure 3B)**.

**Table 1:**
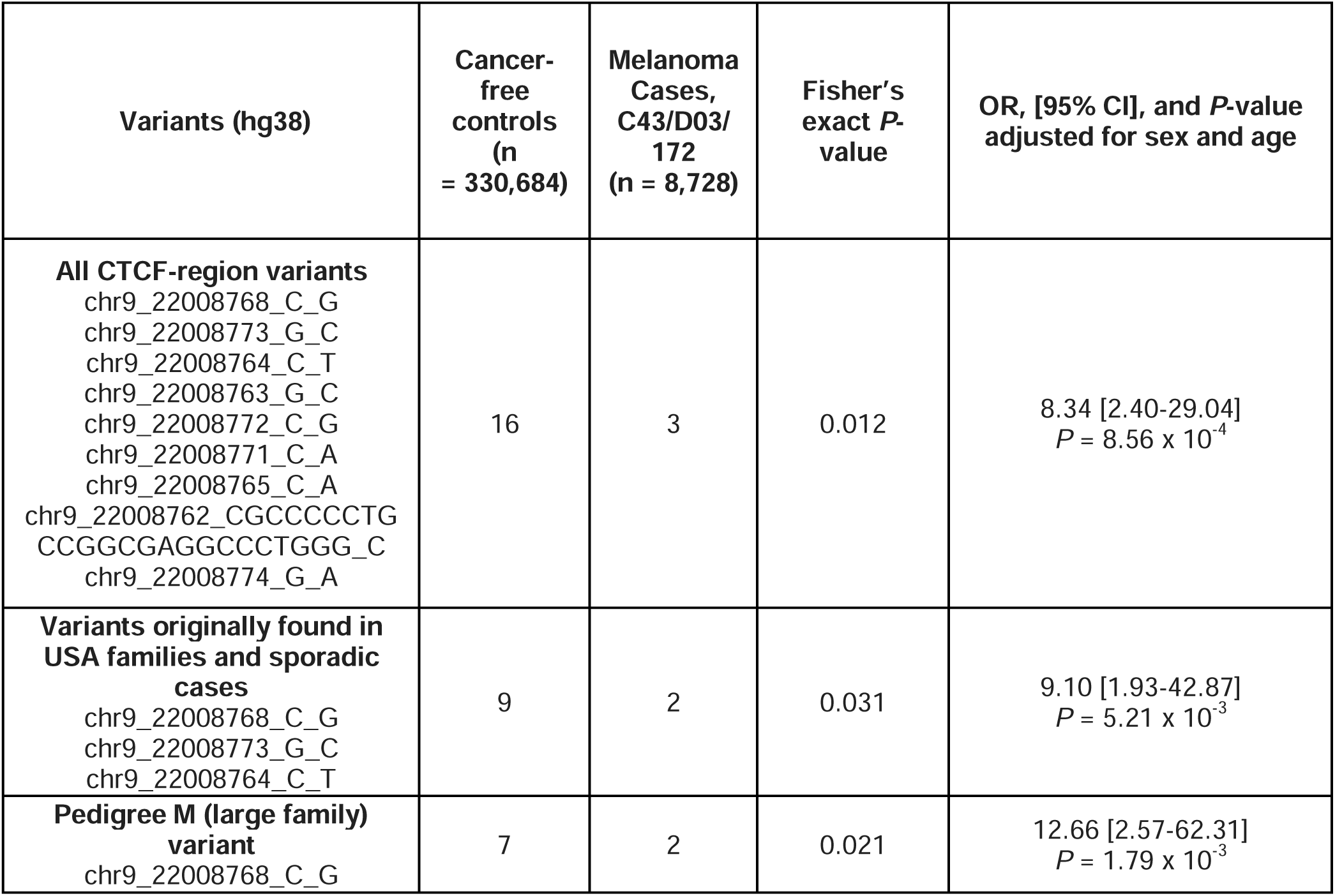
Associations of *CDKN2B* CTCF-motif variants with melanoma risk in the UK Biobank.

### Risk-associated variants disrupt CTCF binding

To validate the impact of the variants on CTCF binding, we first performed an oligonucleotide pulldown assay using nuclear extract purified from immortalized C283T melanocytes, followed by Western blotting with an antibody against CTCF. Compared to the oligo carrying the reference sequence, oligos containing each of the four variants found in American pedigrees or UK Biobank melanoma cases showed diminished binding of CTCF *in vitro* (0.19 to 0.56 relative to the reference sequence; **Figure 3D**). To further dissect the binding of these variants to CTCF in cells, immortalized dermal fibroblasts from the large pedigree M, including an affected carrier and an unaffected carrier (M-A-27 and M-A-32, respectively) as well as an unaffected individual wild-type (WT, M-A-26) for the respective *CDKN2B* variants, were cultured and assayed using CTCF ChIP-seq. CTCF ChIP samples showed significant enrichment of predicted CTCF motifs within ChIP-seq peaks genome-wide compared to the input sample (**Supplementary Figure 5**), indicating that the CTCF ChIP procedure was successful. CTCF ChIP-seq libraries had a clear peak at the *CDKN2B* CTCF binding site containing the variant (chr9:22008758-22008777, hg38) (**Figure 3E**). We then quantified the mutant and WT allele frequencies from the ChIP and input samples for the two variant carriers. For the sample from a carrier melanoma case (M-A-27), the variant allele was detected in only 9.5% of reads from the ChIP sample, while we observed no variant allele reads for the ChIP sample from an unaffected carrier (M-A-32; **Figure 3F**).

Combined, the variant allele frequency (VAF) for the ChIP samples was significantly lower in ChIP samples (VAF=0.025) relative to the genomic DNA input (VAF=0.375, *P* = 0.0002, Fisher’s exact test), suggesting the variant found in pedigree M causes a strong loss of CTCF-binding and indicating functionality of the variant.

CTCF is a repressive transcription factor when bound in solitude, however, two distal bound CTCF sites can be abridged when interacting with the cohesin complex, resulting in a DNA loop. Interestingly, based on previously published High-throughput Chromosome Conformation Capture (HiC) data [63], the region harboring the *CDK2NB* CTCF motif can form a loop with a CTCF binding site in the *MTAP* gene with the resulting DNA loop containing the *CDKN2A* gene (**Supplementary Figure 6**). We then investigated whether this loss of CTCF binding resulted in a loss of the known DNA loop between *CDKN2B* and *MTAP* using high-resolution HiC on the fibroblasts from two carriers from large pedigree M. HiC data analysis was able to confirm the loop between CTCF binding sites of *CDKN2B* and *MTAP* (**Figure 4; Supplementary Figure 6**).

**Figure 4.**
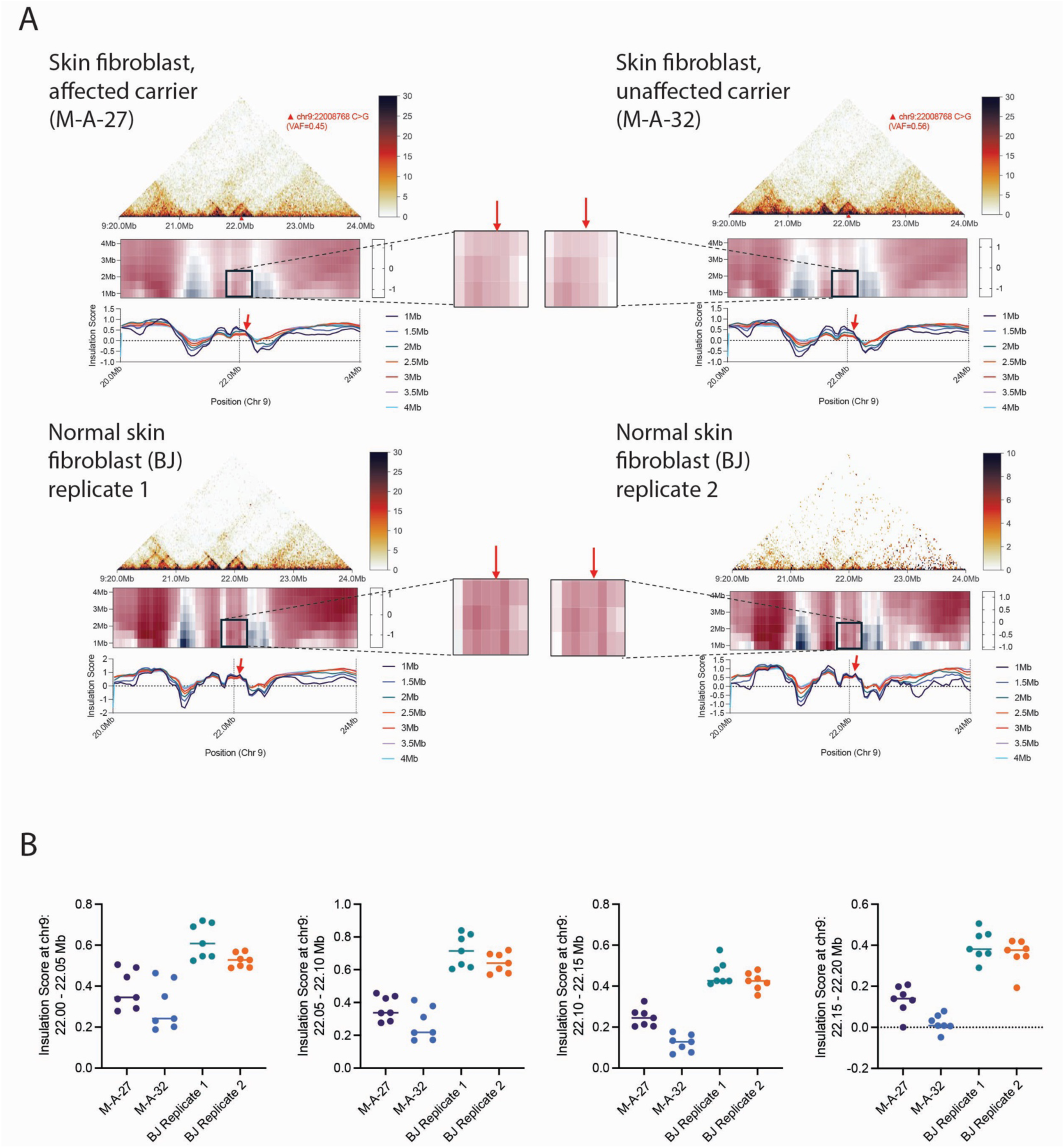
HiC analysis of the *CDKN2A*/*B* locus in immortalized dermal fibroblasts from patients with chr9 g.22008768 C>G (hg38). (**A**) Hi-C contact maps for *CDKN2B* heterozygous fibroblasts from carriers M-1007 (left upper) and M-1012 (right upper) and wild-type BJ cells (lower panels) are shown for chr9: 20-24 Mb. The red triangle marks the *CDKN2B* variant (chr9 g.22008768C>G) with its corresponding variant allele frequency (VAF) indicated for samples M-A-27 and M-A-32. Insulation score profiles based on different window sizes (1 Mb to 4 Mb in 0.5Mb steps, plotted on the Y-axis) are plotted across the genomic region in 50Kb intervals in the middle panel. A zoomed-in view of regions upstream and downstream of the variant (from 21.80 to 22.20 Mb) is shown, with a red arrow highlighting the position near the variant, indicating changes in chromatin insulation at this locus. The same insulation scores are represented by colored curves in the bottom panel with the red arrow pointing to the changes in insulation score. (**B**) Insulation scores for 50 Kb regions spanning from chr9: 22.0 to 22.2 Mb are plotted; individual points represent individual insulation scores calculated from seven windows spanning from 0.5 Mb to 4 Mb in size.

We analyzed the insulation score, which measures the frequency of chromatin interactions crossing a given genomic locus within a defined window size and is commonly used to quantify boundary strength. At the variant locus overlapping the disrupted CTCF binding site, insulation analysis showed a modest reduction in boundary strength in fibroblasts from pedigree M variant carriers (g.22008768C>G) M-A-27 and M-A-32 compared with control fibroblasts (**Figure 4A**), in particular for the region spanning 22.0Mb to 22.2 Mb (**Figure 4B**), indicating weakened local chromatin insulation and reduced boundary strength.

We further examined previously published capture-HiC data generated from human primary melanocytes where the larger 9p21 region was assayed with tiled capture baits [64]. Notably, we observe prominent interactions between the variant region and the CTCF-bound region in *MTAP* consistent with fibroblast data, as well interactions with p14 and p16 promoter regions (**Supplementary Figure 7**), suggesting a potential role for the region harboring these variants in regulation of *CDKN2A* transcripts.

### Familial CDKN2B variants are associated with allele-specific loss of CDKN2A expression

We directly assessed allelic expression of *CDKN2B*, as well as the p16 transcript of *CDKN2A*, in patient-derived fibroblast cell lines from variant carriers. One sequenced carrier from the large M pedigree (M-A-27) is heterozygous for a common 3’-UTR variant (rs11515) within both p14 and p16 *CDKN2A* transcripts, as well as two carriers heterozygous for rs3217992 in the 3’-UTR of *CDKN2B* (M-A-27 and M-A-32). The alleles of these two SNPs residing on the variant haplotype from pedigree M were resolved using long-read whole-genome sequencing (**Supplementary Figure 8**), allowing for assessment of phase-resolved allele-specific expression (ASE). Using RNA-seq data obtained from immortalized dermal fibroblasts, we calculated VAF as a fraction for both rs11515 (*CDKN2A*) and rs3217992 (*CDKN2B* 3’UTR) in samples WT and carriers of g.22008768C>G. Cells were either vehicle-treated or treated with TGF-β to induce *CDKN2B* expression. We compared a variant carrier (case, M-A-27) harboring g.22008768C>G and a married-in non-carrier control (M-A-26), where both are heterozygous for rs11515. The carrier showed lesser *CDKN2A* expression from the haplotype carrying g.22008768C>G, with the VAF differing significantly from that derived from the non-carrier in both vehicle (carrier average VAF = 0.13, range 0.11-0.17; non-carrier average VAF = 0.42, range 0.39-0.48; *P* = 0.0012, unpaired two-tailed t-test) and TGF-β conditions (carrier average VAF = 0.14, range 0.064-0.21; control average VAF = 0.44, range 0.42-0.52; *P* = 0.0069, unpaired two-tailed t-test; **Figure 5A**). For the rs3217992 (*CDKN2B* 3’UTR) variant, most untreated samples did not contain 10 reads, thus, we only considered TGF-β treated samples. Of samples heterozygous for rs3217992, we observed no difference in VAF between variant carriers (M-A-27, M-A-32) and a non-carrier control (M-C-22; mean carrier VAF = 0.60 for the haplotype carrying g.22008768C>G, range = 0.30 - 0.73; non-carrier mean VAF = 0.44, range = 0.32 - 0.68; *P* = 0.32, Welch-corrected unpaired two-tailed t-test) (**Figure 5B**).

**Figure 5.**
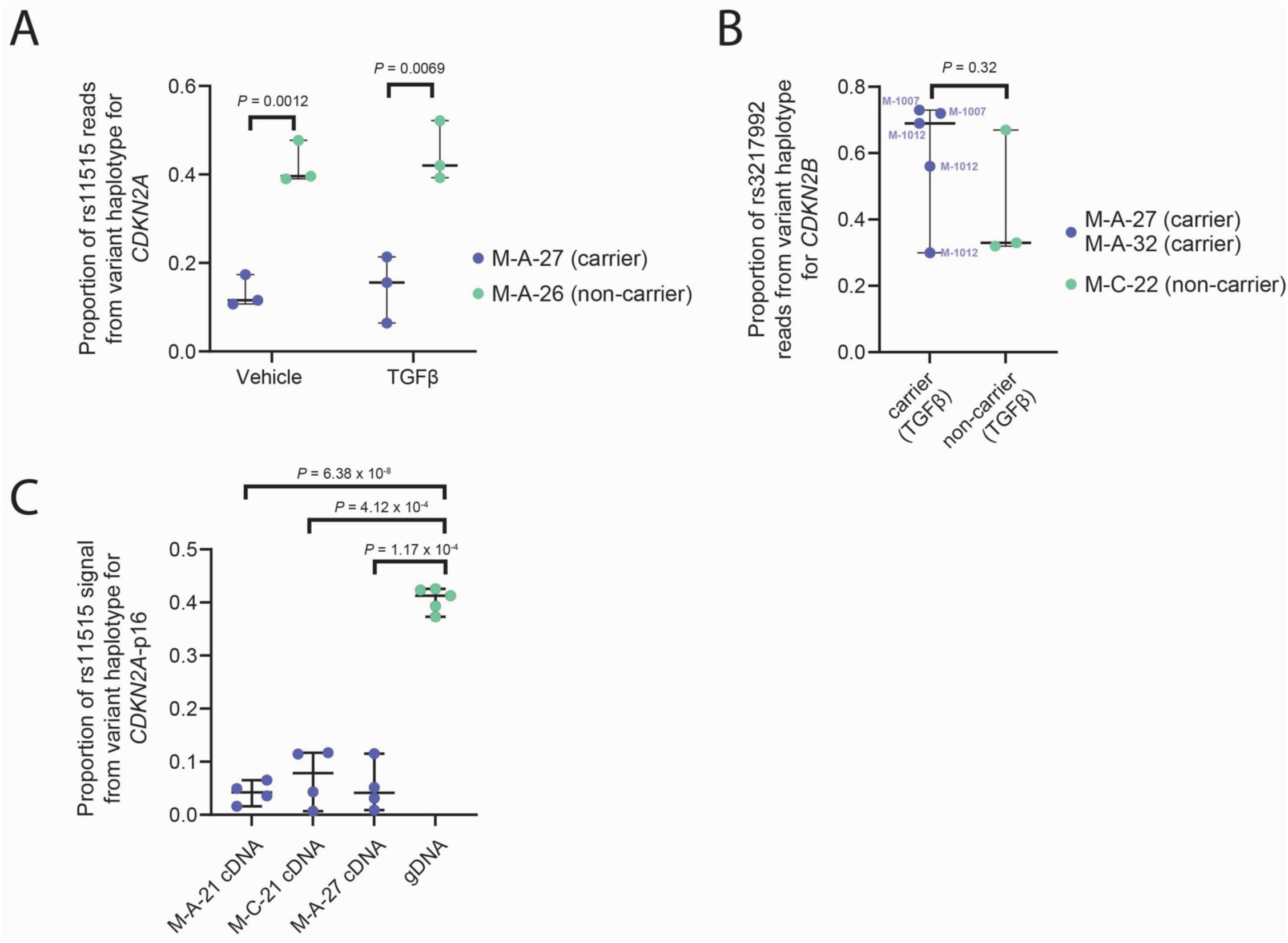
Allele specific expression of p16 (*CDKN2A*) and *CDKN2B* in fibroblasts cell lines from carriers of *CDKN2B* variants in pedigree M. (**A**) Allele specific expression of *CDKN2A* was assessed in fibroblasts from a pedigree M carrier (M-A-27) and non-carrier (M-A-26) heterozygous for CDKN2A 3’-UTR SNP rs11515 with and without TGF-B treatment followed by RNA sequencing. The proportion of allelic reads from the haplotype shared with g.22008768C>G is plotted for three biological replicates for each sample/condition, with differences assessed by unpaired two-tailed t-tests. Values represent reads from p16 and p14 isoforms combined. (**B**) Allele-specific expression of *CDKN2B* was assessed in fibroblasts from two pedigree M carriers and one non-carrier heterozygous for rs3217992 from RNA sequencing data derived following TGF-B treatment. The proportion of allelic reads from the haplotype shared with g.22008768C>G is plotted with differences between carriers (two replicates of M-A-27 and three replicates of M-A-32 combined) and a non-carrier (three replicates of M-C-22) assessed by a Welch-corrected unpaired two-tailed t-test. (**C**) Allele-specific expression of the p16 isoform of *CDKN2A* was assessed in fibroblasts from three pedigree M carriers heterozygous for rs11515 using a Sanger-sequencing based isoform-specific assay. The proportion of allelic signal (peak height) from the haplotype shared with g.22008768C>G is plotted for four biological replicates of each patient fibroblast, with differences assessed by Welch-corrected unpaired two-tailed t-tests relative to heterozygous genomic DNA (five replicates). Blue dots represent biological replicates of cDNA from three patients and green dots represent gDNA samples. All plots are shown as median with range.

While we observe allele-specific expression in patient fibroblasts, short-read RNA sequencing cannot distinguish between expression from the p16 and p14 *CDKN2A* transcripts. We therefore assessed allelic expression of p16 in fibroblasts from patient carriers using a p16-specific Sanger sequencing-based assay, observing little-to-no p16 expression driven from the haplotype carrying g.22008768C>G in all three carrier cases heterozygous for rs11515 that we assayed (*P* = 6.38 x 10^-8^, *P* = 4.12 x 10^-4^, and *P* = 1.17 x 10^-4^ for individuals M-A-21, M-C-21, and M-A-27, respectively relative to heterozygous genomic DNA; Welch-corrected unpaired two-tailed t-test; **Figure 5C**). We were unable to assess p16 allelic expression in samples from pedigree X as none were heterozygous for the rs11515 *CDKN2A* proxy SNP.

### Perturbation of the CDKN2B CTCF binding site affects CDKN2A gene expression in cultured melanocytes

We next sought to assess *cis*-regulatory function of the region harboring melanoma-associated risk variants using CRISPR-inhibition (CRISPRi). Specifically, we targeted the region of the CTCF-motif simultaneously with two small guide RNAs (sgRNAs) in both primary (C87) and immortalized human melanocytes (C283T) and assessed the expression of genes within the 9p21 region. In immortalized melanocytes, relative to non-targeting control sgRNAs, the region-specific sgRNAs broadly disrupted expression of multiple transcripts in the 9p21 region, including increased expression of *DMRTA1* (1.78-fold; *P* = 0.0001), while reducing expression of *MTAP* (0.64-fold; *P* = 0.035), *CDKN2A-AS1* (0.68-fold; *P* = 0.016), and critically reducing both the p16 and p14 transcripts of *CDKN2A* (0.71-fold and 0.66-fold, *P* = 0.0022 and *P* = 0.011, respectively; **Figure 6A**); *CDKN2B* expression was not significantly changed (*P* = 0.33). We observed a mostly similar trend in primary melanocytes with significant decreases in *MTAP* (0.62-fold, *P* = 0.0060), *CDKN2B-AS1* (0.49-fold, *P* = 0.0024), and both p16- and p14-*CDKN2A* (0.74-fold and 0.56-fold, *P* = 0.049 and *P* = 0.015, respectively), but not *CDKN2B* (*P* = 0.37; Figure 6B).

**Figure 6.**
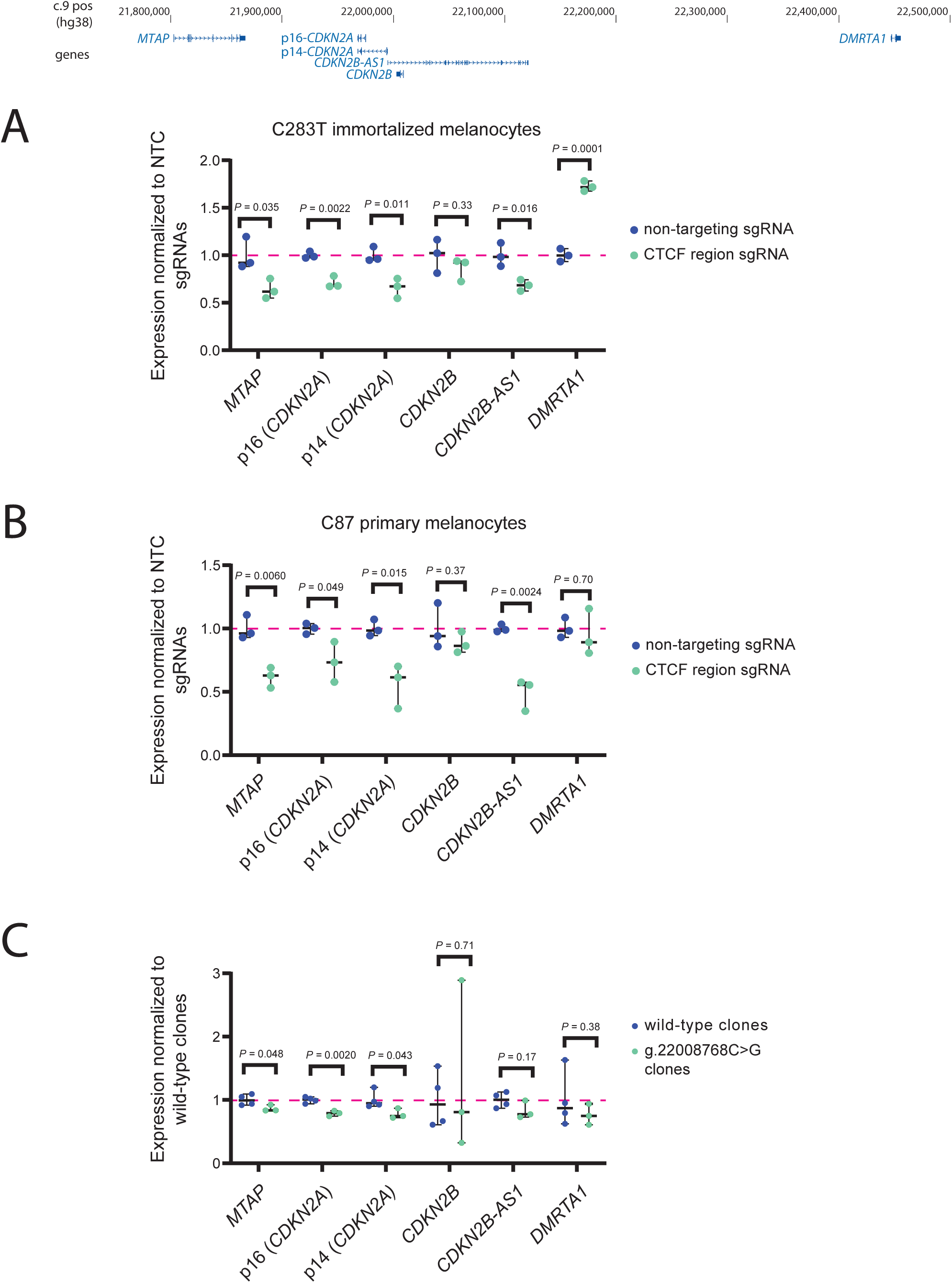
Effect of CRISPR-interference and CRISPR-editing of variants on expression of genes within the 9p21 region in melanocytes. CRISPR-inhibition (CRISPRi) was performed to target the region harboring familial *CDKN2B* variants and assess potential *cis*-regulatory targets in (**A**) C283T immortalized melanocytes and (**B**) C87 primary human melanocyte cultures. Two small guide RNAs (sgRNAs) were designed to target the region surrounding familial variants. C283T cells stably expressing dCas9-KRAB were infected with lentiviral particles expressing both sgRNAs. For C87, cells were co-infected with lentiviral particles expressing sgRNAs and expressing dCas9-KRAB, respectively. Expression of transcripts in the 9p.21 region relative to the expression of *TBP* was measured via TaqMan quantitative RT-PCR assays and further compared to expression from cells instead targeted with two non-targeting sgRNAs. Each data point represents the average of three technical replicates for each of three biological replicates. Plot shows expression levels relative to the average of the non-targeting guide biological replicates with median with range. *P*-value was calculated using unpaired two-tailed t-tests. (**C**) 9p21-region gene expression in genome-edited immortalized melanocytes (C283Ts) heterozygous for the chr9 g.22008768C>G (pedigree M) variant. Three clones heterozygous for g.22008768C>G were compared to four clones homozygous for the reference allele; gene expression was quantitated using TaqMan quantitative RT-PCR assay showing of genes in the region relative to *TBP*. Each data point represents the average of three technical replicates of four biological replicates for each unique clone. Plot is shown as expression normalized to the average of that from wild-type clones as median with range. *P*-value was calculated using Welch corrected unpaired two-tailed t-test. The location of genes tested within the 9p21 region is shown in the upper panel.

We next sought to evaluate the effect of familial variants on expression of 9p21 genes. Specifically, we edited the g.22008768C>G variant from the larger M pedigree into C283T immortalized melanocytes, obtaining three independent clones heterozygous for the variant the (full sequence analysis of wild-type and heterozygous clones at the sgRNA site is provided in the **Methods and Supplementary Figure 9**). Consistent with the CRISPRi data, we observed significant differences of expression for multiple transcripts, with reduced expression in edited clones relative to wild-type clones for *MTAP* (0.86-fold, *P* = 0.048), p16-*CDKN2A* (0.79-fold, *P* = 0.0020), and p14-*CDKN2A* (0.78-fold, *P* = 0.043), but not *CDKN2B* (*P* = 0.71; **Figure 6B**). In summary, both CRISPR-interference and CRISPR-editing of a variant disrupting CTCF binding in cultured melanocytes alters the expression of multiple transcripts in 9p21 region, critically including the p16 and p14 transcripts of *CDKN2A*.

## Discussion

While *CDKN2A* was identified as the major melanoma susceptibility gene more than three decades ago [5, 65], a proportion of families linked to the 9p21 locus remain unexplained by protein-coding *CDKN2A* variants. Despite the search for pre-disposing non-coding variation within *CDKN2A* [27] or risk-associated variation in other genes at this locus, most such families remain genetically unexplained. A number of targeted studies of high-risk families have investigated *CDKN2B* as a candidate [23, 24, 42–45], however, rare protein-coding *CDKN2B* variants have never been established to be associated with risk. Here, we report rare variants initially identified as protein-coding variants in a non-canonical isoform of *CDKN2B* and show that, while they are associated with risk, they appear to do so at least in part via cis-regulation of the established gene at the locus, *CDKN2A*.

Of the three variants we identified from WES data from melanoma families and a case-control study, all three are missense for an alternative transcript of *CDKN2B*, p10 [51], generated from an alternative 5’ splice donor site in intron 1. While the N-terminal end of p10 is identical [57] to that of p15 (Ink4b), and like p15, is induced by transforming growth factor beta, it does not appear to form a complex with CDK4 or CDK6 [51], suppress tumorigenicity in glioblastoma cell lines [52], nor does it interfere with the p15 association with these two cyclin-dependent kinases [51]. Our analysis of junction reads in melanocytic cells, however, suggest that most melanocytes and melanomas express little to no p10 transcript, so while these three variants are predicted to impact p10 function, the lack of p10 expression argues against these variants functioning significantly through p10. On the other hand, all three identified variants are located in an intronic CTCF-binding motif of p15, a transcript shown to induce cell cycle arrest in mouse models [53] and is highly expressed in melanocytes and melanoma cell lines. Notably, while the sample numbers are small, the rare novel CTCF-motif variant we found in a melanoma case from the UK Biobank is not a missense variant for p10, but rather, like the other three variants, was predicted to have a strong effect on CTCF-binding (**Supplementary Table 4**). These findings reinforce the interpretation that altered chromatin regulation, rather than changed protein function, underlies the disease risk.

While attention has increasingly focused on functional non-coding variants, predicting and validating their effects remains challenging. Google DeepMind recently developed AlphaGenome [57], a regulatory variant effect prediction tool which offers promise. Our study provides a compelling use case: AlphaGenome predicted loss of CTCF binding for all three variants we identified in American melanoma cases (**Figures 3A-B**), which we experimentally validated through ChIP-seq. Via previously published and new chromatin conformation-based data from dermal fibroblasts, we were able to show that this motif is located within a prominent chromatin loop from *CDKN2B* to the *MTAP* gene, with the variant observed in our larger pedigree resulting in weaker chromatin interactions as evidenced by reduction in insulation score in carrier fibroblasts. Using high-resolution capture-HiC, we also show that this region physically interacts with annotated promoter sequences over the p16 and p14 transcripts in melanocytes, the cell type of origin for melanoma. The broader mechanism by which loss of this loop results in altered expression of 9p21 genes remains to be established, however, CRISPR-inhibition and CRISPR editing data suggest effects on more than one gene within the region.

Multiple lines of evidence collectively point to these variants functioning at least in part through *CDKN2A*: allele-specific expression of p16 in patient fibroblasts, reduction of p16 and 14 expression following targeting of this region using CRISPR-inhibition, and reduction of expression of p16 and p14 in melanocyte clones edited to carry g.22008768C>G. We also note that broadly, the cases from both pedigrees had phenotypes that are hallmarks of families carrying pathogenic variants in *CDKN2A*: early age of onset, multiple primary melanomas, dysplastic nevi, and a high number of nevi [7, 9, 25, 66]. While our work does not exclude a potential role for other genes in the region and indeed suggests that these variants may have broader *cis*-regulatory effects via loss of CTCF-binding, it provides strong evidence for function of these variants through loss of p16 and/or p14.

Disruption of CTCF-mediated chromatin regulation has been increasingly implicated in cancer, but predominantly in the somatic setting. For example, somatic deletions of CTCF binding sites result in loss of topological insulation and oncogene activation (e.g., TAL1 or LMO2) in T-cell acute lymphoblastic leukemia [67]. Similarly, disruption of CTCF insulators in mouse glioma models leads to PDGFRA activation and tumorigenesis, accompanied by loss of *CDKN2A* expression through promoter hypermethylation [68]. In contrast, the role of germline CTCF disruption in cancer susceptibility—particularly through point variants affecting core CTCF binding motifs—has remained largely unexplored. To our knowledge, this study represents one of the first demonstrations that inherited sequence variants disrupting a CTCF binding site can act as a high-risk susceptibility mechanism for familial cancer by altering tumor suppressor gene expression. These findings reveal a previously unrecognized mechanism of non-coding *cis*-regulatory variation affecting a well-established melanoma susceptibility gene and suggest that other variants in this region with broad-regulatory effects could confer risk and potentially explain additional 9p21-linked *CDKN2A* mutation-negative families, possibly through loss of CTCF binding.

Our study has several limitations. First, the number of families in which these variants were identified is small, although one of the families includes more than ten melanoma cases spanning multiple generations. The extremely low allele frequency of these variants also limited statistical power to assess melanoma risk associated with these variants even in large biobanks, although we observed significant enrichment of melanoma cases among carriers of the pedigree M variant, pedigree M and X variants, or for that matter any variant overlapping the core motif sequence in UK Biobank. Second, the absence of tumor tissue from variant carriers has precluded evaluation of somatic second-hit events, such as LOH or promoter hypermethylation of 9p21 genes, which may act in tandem with CTCF disruption in a single allele.

In summary, we identified a novel susceptibility mechanism for a well-known melanoma predisposition gene in two families linked to 9p21 in which prior efforts failed to identify a causal gene. Our findings support the hypothesis that non-coding regulatory variants can function as high-penetrance susceptibility alleles and contribute to missing heritability in familial melanoma. More broadly, they highlight the importance of considering disruption of CTCF-binding when evaluating unresolved cancer-prone families and suggest that systematic interrogation of CTCF binding sites at known cancer predisposition loci may uncover additional cancer susceptibility genes.

## Methods

### Whole exome and panel sequencing of familial and sporadic melanoma cases

Whole-exome sequencing (WES) of high-risk melanoma families from the United States and population-based controls from Cancer Prevention Study-II was performed as previously described [46, 47], as was gene panel sequencing of melanoma cases and controls drawn from the Prostate, Lung, Colon, Ovary (PLCO) screening trial [47]. The NCI/NIH Familial Melanoma Study (Clinical Trials Number: NCT00040352) was approved by the NIH IRB and all participants provided written informed consent. Informatic analysis has also been previously described [19, 47, 69, 70]. Genotype calling was performed using three different variant callers (HaplotypeCaller and UnifiedGenotyper from GATK, as well as FreeBayes 9.9.2) including all target regions +/- 250 bp flanking sequence. An Ensemble variant calling pipeline (v0.2.2) integrated genotype calling results and applied a Support Vector Machine algorithm to finalize genotype calls. Insertions and deletions were left-aligned post-alignment (BAM) as well as post-variant-calling (VCF) via GATK’s LeftAlignIndels and LeftAlignVariants modules.

### Variant annotation

Variants were annotated for predicted protein-coding function using annovar [71] (https://annovar.openbioinformatics.org/en/latest/) and for predicted alteration of sequence-specific protein binding using motifbreakR [58, 59] (https://github.com/Simon-Coetzee/motifBreakR).

Regulatory effects of the identified variants were assessed using the deep learning framework AlphaGenome [57] as implemented in Google Colab. Analyses were performed on the human reference genome (hg38). Each variant was modeled by comparing WT and mutant alleles within a 1 Mb genomic window centered on the variant position. Predictions were generated in a dermal fibroblast context (Cell Ontology: CL:0002551) for transcription (RNA-seq), chromatin accessibility (DNase I hypersensitivity), CTCF binding (CTCF ChIP-seq), and active enhancer activity (H3K27ac ChIP-seq). Predicted signal tracks for WT and mutant alleles were extracted and visualized to identify variant-associated changes in regulatory activity across the locus.

### Long Read (PacBio) whole genome sequencing of family members and immortalized melanocytes

Long-read whole-genome sequencing (WGS) sequencing was performed using genomic DNA extracted from fibroblasts of two members of Family M (A-21 and C-21, both are affected carriers), and from C283T parental cells from which family M variant HT clones were generated. Sequencing was performed as described in the manufacturer’s protocol for “Preparing whole genome and metagenome libraries using the SMRTbell® prep kit 3.0” (PacBio, Menlo Park, CA, USA, 102-166-600), with modifications. Briefly, 1.25 µg of genomic DNA (for the AMPure size-selection workflow) or 3 µg of genomic DNA (for the PippinHT size-selection workflow) was used as an input, determined by Quant-iT™ PicoGreen dsDNA Reagent (ThermoFisher Scientific, Waltham, MA, USA). DNA was sheared to a target fragment size of 15-20 kb using the FastPrep-96 (MP Biomedicals, LLC, Solon, OH, USA).The resulting fragmented DNA had damage and end repair performed, ends were A-tailed, and barcoded overhang adapters were ligated to the fragment ends to generate SMRTbell libraries, followed by nuclease treatment to remove unligated fragments. For AMPure-based size selection, libraries underwent a stringent AMPure PB bead purification to remove small fragments (<5 kb). For Gel-based size selection, fragments smaller than 10 kb were removed using the PippinHT instrument (Sage Science, Beverly, MA, USA), with a 1x SMRTbell cleanup performed both before and after size selection. Following purification, the primer annealing and polymerase binding of the SMRTbell libraries were performed according to PacBio’s protocols. The prepared SMRTbell libraries were then sequenced on the PacBio Revio instrument using 25M SMRT Cells. CCS (circular consensus sequencing) reads were generated using SMRT Link v11. Reads were aligned to hg38 using pbmm2. Small variant calling was performed using DeepVariant [72] and GATK HaplotypeCaller [73], and phasing of these variants was performed using WhatsHap [74]. Structural variants were called using cuteSV [75, 76] and pbsv (https://github.com/PacificBiosciences/pbsv).

### Identification of CDKN2B Isoform Expression in Melanocytes & Melanoma

To quantify the expression of *CDKN2B* isoforms in melanocytes and melanoma cell lines, RNA-seq data were analyzed from 106 human primary melanocyte cultures [55], 75 melanoma cell lines [56], and 445 tumors from the TCGA-SKCM melanoma dataset [30]. Read alignment was performed as previously described. We extracted junction-level information from each dataset, including specific splice junctions and the number of supporting reads. Exon–exon junction counts corresponding to *CDKN2B* transcripts were processed, and data for the two junctions of interest (chr9:22,006,246–22,008,673, p10-*CDKN2B*, NM_078487; chr9:22,006,246–22,008,796, p15-*CDKN2B*, NM_004936; hg19) were summarized (**Supplementary Table 3**). Junction usage was normalized using the ratio of junction read depth to the total read depth on the 3’ end (relative to the *CDKN2B* transcript) of the junction sites for visualization.

### Cell culture and treatments

Immortalized patient fibroblasts from family members harboring germline pathogenic variants as well as unaffected non-carriers were cultured in IMDM supplemented with 20% FBS, 1x glutamine and penicillin/streptomycin at 5% CO_2_. Frozen aliquots of melanocytes were isolated from the foreskin of healthy newborn males, mainly of European descent, following an established protocol from the Specialized Programs of Research Excellence (SPORE) in Skin Cancer Specimen Resource Core at Yale University, were obtained. Cells were either grown in Dermal Cell Basal Medium (American Type Culture Collection/ATCC PCS-200-030) supplemented with Melanocyte Growth Kit (ATCC PCS-200-041) and 1% Amphotericin B/Penicillin/Streptomycin (Quality Biological, Cat# 120-096-711) or in Gibco Cascade Biologics M254 (Invitrogen, Cat# M-254-500) supplemented with HMGS-2 (Invitrogen, Cat#S0165), grown at 37°C with 5% CO_2_. All cells tested negative for mycoplasma via the MycoAlert PLUS Mycoplasma Detection Kit (Lonza, Cat# LT07-710). TERT-immortalized melanocytes [77] were cultured in the same manner.

### CTCF pulldown by oligonucleotides carrying non-coding variants

Biotin-labelled annealed double-stranded oligonucleotides (450pmol) were immobilized on 30 ul neutravidin beads (Cat#29200, Thermo Scientific, CA) or 2mg Dynabeads Myone Dtreptavidin T1 beads(Cat# 65601, ThermoScientific) in DNA binding buffer (50mM HEPES, pH 8.0, 150 mM NaCl), followed by 3x washes in 1 ml DNA binding buffer to remove unbound oligonucleotides. The immobilized oligonucleotides were incubated with 130 ug nuclear extract purified from C283T melanocytes in the presence of 10 ug poly(dI-dC) (Cat#20148E, Thermo Scientific, CA) for 2 hours at 4°C with constant rotation. The beads were washed three times with DNA binding buffer and resuspended in 2x LDS solution (Thermo Scientific, CA) supplemented with reducing agent. The beads were boiled and the supernatant was resolved on a 4-12% NuPAGE Bis-tris gel (Thermo Scientific, CA). Western analysis was performed using rabbit monoclonal CTCF antibody at 1:1000 dilution (Cell Signaling, MA). The gel-image was analyzed and band volume was quantitated via local background using Image Lab software by Bio-Rad. Oligonucleotide sequences are included in **Supplemental Table 5**.

### RNA sequencing

Fibroblasts from four members of pedigree M were sequenced, including two carriers (M-A-27, melanoma case; and M-A-32 carrier with a dysplastic nevus diagnosis) and two controls (M-A-26 and M-C-22). Cells were seeded at 20,000/cm^2^ into 60 mm TC dishes and allowed to grow to ∼80% confluence for ∼48 hours before being treated with 200 nM TGF-β(R&D Systems, 240-B-002) or vehicle for 8 hours. RNA extraction was performed using QIAGEN RNeasy mini kit following manufacturers protocol for RNA extraction of animal cells. RNA was submitted to Novogene for library preparation and sequencing. Stranded, polyA-based library preparation was used and libraries were sequenced as 150 base, paired-end reads at approximately 25 million read pairs per library on Illumina NextSeq 500 or Illumina NextSeq 2000. Three biological replicates were performed for each culture/condition. Fastq files were aligned to UCSC hg38 using STAR [78] with hg38.ensGene.gtf to specify splice junctions. Reads for each allele of rs11515 (*CDKN2A*) and rs3217992 (*CDKN2B*) were counted from bam files using bam-readcount [79] for those samples heterozygous at these positions for replicates with at least 10 reads over the respective position. Variant allele frequency (VAF) of the allele(s) residing on the same haplotype as g.22008768C>G (rs11515-C and rs3217992-C, oriented to the plus strand of the genome) was counted as a simple fraction of mutant over total read counts and the significance of difference was tested via and unpaired two-tailed t-test and Welch corrected unpaired two-tailed t-test for alleles of rs11515 and rs3217992, respectively. Three replicates each passed quality control with the exception of M-A-27 treated with TGF-β, for which only two replicates were usable.

### Chromatin Immunoprecipitation Sequencing (ChIP-seq)

CTCF chromatin immunoprecipitation coupled with next generation sequencing (ChIP-seq) was performed essentially as described [80, 81]. 20 x10^6^ cells grown in 150 mm dishes were fixed in 1% formaldehyde PBS for 10 minutes at room temperature then quenched in 125 mM glycine. Fixed cells were washed in ice-cold PBS then scraped in PBS and resuspended gently in ice-cold lysis buffer (10mM Tris pH8.0, 10mM NaCl, 0.2% NP40, supplemented with protease and phosphatase inhibitors). Samples were incubated on ice for 10 minutes to assist lysis then pelleted by centrifugation before being snap frozen in liquid nitrogen and then stored at −80°C. Pellets were thawed on ice and resuspend in nucleic lysis buffer (50mM Tris pH8.0, 10mM EDTA pH8.0, 1% SDS, supplemented with protease and phosphatase inhibitors), then sonicated using the Diagenode Bioruptor Pico at 4°C 10 cycles of 30 seconds on, 30 seconds off. Protein G dynabeads (Invitrogen 10003D) were coupled to IgG (Sigma I5006) or anti-CTCF (Millipore 07-729) by washing beads in cold PBS then incubating in PBS overnight with 10 µg of antibody with rotation.

Sonicated chromatin was pre-cleared by incubating with the IgG coupled beads for 1 hour at 4°C with rotation. The supernatant was retained and incubated with anti-CTCF overnight at 4°C with rotation. CTCF immunoprecipitated chromatin was recovered by collecting protein G dynabeads on a magnetic rack and washing twice with salt wash buffer (20mM Tris pH8.0, 2mM EDTA, 50mM NaCl, 1% TritonX100, 0.1% SDS), 1 x with lithium chloride wash buffer (10mM Tris pH8.0, 1mM EDTA, 0.25M LiCl, 1% NP40, 1% sodium deoxycholate) then twice with Tris-EDTA buffer (10mM Tris pH 8.0, 1mM EDTA). Immunoprecipitated DNA was then eluted from the beads by incubating in 100mM NaHCO_3_, 1% SDS for 30 minutes at 65°C with occasional vortexing. RNase A (Invitrogen, EN0531) digest was performed then cross-links were reversed by incubation in 0.3M NaCl at 67°C overnight. The next morning, 60 µg proteinase K (Invitrogen, 25530049) was added to digest cross-linked proteins at 45°C for 2 hours and finally, DNA was purified using a phenol-chloroform DNA extraction. Immunoprecipitated and input DNA were sent to BGI (Hong Kong) for library preparation and sequencing on a BGISEQ500. ChIP libraries were sequenced as 50 base single-end reads while the input (WGS) library was sequenced as 100 base, paired end reads.

Fastq files were aligned to UCSC hg38 using bwa-mem2 [82] then converted from sam to bam file with samtools view [83]. Duplicates were marked with picard (https://broadinstitute.github.io/picard/) MarkDuplicates and subsequently removed using samtools view [83]. Deeptools [84] was used to profile input-normalized CTCF ChIP-seq coverage around the CTCF motif center of ∼31,000 CTCF-Cohesin binding sites obtained from supplementary materials of Katainen *et al*. [85] and lifted over to hg38. Briefly, bam files were converted to bigwig using bamCoverage with RPGC normalization then CTCF ChIP and IgG were normalized to Input using bigwigCompare. Matrices were built with a 2000 base pair flank either side of the center of CTCF motifs in 10 base pair bins and plotted. Variant and WT reads were calculated on deduplicated bam files using bam-readcount [79]. We performed this count on whole CTCF motif harboring the variant (chr9:22,008,758-22,008,777 in hg38) and the results were parsed in R.

### High-throughput chromosome conformation capture (HiC)

HiC was performed essentially as described previously for adherent cells (in the supplementary materials) [86]. 5 x 10^6^ cells were fixed in serum free Hand’s Balanced Salt Solution (HBSS) and cross-linked with 1% formaldehyde (final concentration) for 10 minutes at room temperature and the reaction was subsequently quenched with 125 mM glycine. Cells were scraped, washed with PBS and pelleted before being snap frozen in liquid nitrogen and stored at −80°C. Cells pellets were thawed on ice and resuspended in 1 mL ice cold lysis buffer (10 mM Tris-HCl, 10 mM NaCl, 0.2% NP-40 supplemented with protease/phosphatase inhibitors). Pellets were incubated on ice for 15 minutes with gentle strokes using a Dounce homogenizer, centrifuged, and then washed with and resuspended in 1x NEB 3.1 buffer. Chromatin was opened by adding SDS to a final concentration of 0.1% followed by incubation at 65°C for 10 minutes. Samples were then chilled on ice and quenched with a final concentration of 1% triton X-100. 10x NEB 3.1 buffer was added to re-adjust the final concentration to 1x, then a restriction enzyme digest was performed with 400 U of DpnII (NEB, R0543L) shaking at 37°C overnight. The enzyme was heat inactivated at 65°C for 20 minutes. To fill in ends with biotinylated ATP, dCTP, dGTP, dTTP and biotin-14-dATP (Thermo Fisher, 19524016) and 50 U DNA polymerase I Klenow were incubated in 1x NEB 3.1 buffer at 23°C for 4 hours shaking.

Ligation was performed using T4 DNA ligase (M0202S) for 16°C for 4 hours with shaking. Fresh ATP was added to the ligation buffer and then the incubation was continued overnight. Cross-links were reversed by digesting the sample with 0.5 mg proteinase K (Invitrogen, 25530049) digestion and incubating at 65°C overnight. HiC DNA was recovered by phenol-chloroform extraction followed by RNase (Invitrogen, EN0531) digest.

To remove biotin from unligated ends, the HiC DNA was incubated with 15U T4 DNA polymerase (NEB, M0203S), dATP, dGTP in NEBuffer 2.1 at 20°C for 4 hours. HiC DNA was then fragmented to 200-300 bp by sonication using the Covaris S2 and fragments were selected by AMPure XP beads (Beckman, A63880). End repair was performed 7.5U T4 DNA polymerase (NEB, M0203S), 25U T4 polynucleotide kinase (NEB, M0201S), 2.5U Klenow DNA polymerase I (NEB, M0210S) and dNTPs 30 min at 20°C, followed by 20 mins at 75°C to deactivate the enzymes. Biotinylated ATP was pulled down using streptavidin beads (Dynabeads MyOne Streptavidin C1, Thermo Fisher 65001) and HiC DNA was then dA-tailed in NEBuffer 2.1 with dATP and 15U Klenow fragment (3’ → 5’ exo-) (NEB, M0212S) for 37°C for 30 minutes and then inactivated at 65°C for 20 minutes on beads, and lastly recovered on a magnetic rack.

Illumina adapters (NEBNext multiplex oligos for illumina, NEB E7335S) were ligated following the manufacture’s protocol and libraries were amplified by PCR (NEB next Ultra II Q5 Master Mix, NEB M0544L). Final libraries were recovered from AMPure XP beads and were stored at - 80°C until sequencing at Novogene (China), when they were sequenced as 150 bases paired-end reads on the Illumnina HiSeq at a total of ∼165M read pairs per library.

### HiC data analysis

The Hi-C sequencing data were processed utilizing HiC-Pro (version 3.1.0) for downstream analysis. Reads were mapped against the human reference genome UCSC hg38 with the use of Bowtie2 [87–89] (version 2.4.4), employing default settings for sensitivity (--bowtie2 --very-sensitive). Quality control measures included setting a minimum mapping quality of 10 (--min_mapq) and filtering for fragment sizes ranging from 50 to 100,000 base pairs (--min_frag_size 50 --max_frag_size 100000) were set to filter out low-quality reads. Valid interactions were filtered based on the ligation site of DpnII fragments and creating contact maps at 40 kb, 150 kb, 500 kb, and 1 Mb resolutions. These matrices were then transformed into .hic format for compatibility with downstream analysis tools using the hicpro2juicebox.sh script provided by HiC-Pro [90]. A wildtype control was established by obtaining replicates of Hi-C data for normal fibroblasts from the ENCODE project (ENCFF155CFR.hic and ENCFF236CFQ.hic). For comparability, all .hic files underwent normalization with the VC_SQRT method via juicer_tools (version 1.9.9; https://github.com/aidenlab/JuicerTools).

Insulation scores were calculated using FANC (version 0.9.28; https://vaquerizaslab.github.io/fanc/index.html) with 1 to 4 Mb window sizes in 0.5 Mb steps. Default settings were applied, including normalization to the chromosomal average and a log2 transformation. For the visualization of region Chr9:20Mb to 24Mb, the contact map was generated using the FANC-plot function and the insulation scores were plotted using GraphPad Prism.

### Melanocyte capture Hi-C

Capture HiC on primary neonatal human melanocyte cultures from five unrelated donors was performed as previously described [64]. The design tiled capture-baits covering all possible restriction fragments of a ∼660 kb region spanning from *MTAP* to past *DMRTA1* on 9p21 (chr9:21,790,755–22,452,478, hg19). For visualization and assessment of chromatin loops given that all interactions were bait-to-bait interactions, we filtered the set of previously analyzed interactions with CHiCAGO score >5 to highlight those with scores greater than the 80^th^ percentile of scores. Melanocyte promoters displayed in **Supplementary Figure 7** are the union of annotated promoter regions from Epigenome RoadMap Project ChromHMM data [91] from two primary melanocyte cultures curated as previously described [64].

### Generation of CRISPR-edited immortalized melanocyte clones for chr9 g.22008768C>G (familyM)

g.22008768C>G (family M) was edited into immortalized human melanocytes (C283T) by homology directed repair (HDR)-based CRISPR editing through electroporation of a ribonucleoprotein (RNP) complex into cells, following procedures as previously described [92] with minor modifications. Immortalized human melanocyte C283T cells were electroporated using the Amaxa P2 Primary Cell Nucleofector kit (X4SP-2096, Lonza) in 20 μL Nucleocuvette strips (Lonza, Cat#V4XP-9096) with the Lonza 4D-Nucleofector Core and X Unit, as recommended by the manufacturer. Briefly, 20 μM or 30 μM SCR7 (Sigma, Cat#D8418-100mL) treated C283T cells were trypsinized, washed, and resuspended in P2 Primary Cell Solution (Lonza, Cat#PBP2-02250) mixed with the provided Supplement 1 solution.

Cas9:sgRNA RNPs were formed by combining an sgRNA targeting a sequence upstream of the chr9:22008768 (120 pmol G2 sgRNA, Integrated DNA Technologies; **Supplementary Table 5**), recombinant Cas9-GFP protein (5 μg, Sigma, Cat#ECAS9GFPPR-250ug), a double-stranded plasmid HDR template harboring the g.22008768-G allele (LFp120; 2 μg). LFp120 was built by cloning homology arms synthesized using Twist into backbone pRC0291 using isothermal assembly. pRC0291 was made by cloning a blasticidin cassette into vector pGMC00018: https://www.addgene.org/195320/. In some electroporations, POLQ siRNA-765 (100 pmol; Integrated DNA Technologies, Cat#593699808, hs.Ri.POLq.13.8, 10 nmol) was included and then electroporated into C283T cells resuspended in P2 buffer using protocol DT130. After electroporation, C283T cells were plated directly into media containing SCR7 to enhance HDR with LFp120. We then isolated monoclonal cell lines from the bulk population through limited dilution plating. For candidate clones grown from single cells, we sequenced the genomic region around chr9:22008768 and deconvoluted the sequences using DECODR (www.decodr.org) to identify potential cell clones carrying the family M variant. We identified three clones heterozygous for g.22008768C>G; each had small insertions/deletions around the cut site on the wild-type allele (**Supplementary Figure 9**) which allowed us to confirm these clones are distinct. As negative controls, we isolated four clones with no change of the sequence at chr9:22008768 (but harbored small insertion/deletions at the cut-site; **Supplementary Figure 9**).

### Quantitative RT-PCR measurement of 9p21 genes in CRISPR-edited melanocyte clones

Gene expression was measured by quantitative real-time PCR (qPCR) using TaqMan Gene Expression Assays (Thermo Fisher Scientific, **Supplementary Table 5**) with standard time conditions (1.6°C/s for: 2 min at 50°C, 10 min at 95°C, and 40 cycles of [15 s at 95°C, 1 min at 60°C]) on an Applied Biosystems QuantStudio 7Flex Real-Time PCR System with QuantStudio Real-Time PCR Software (v1.7.2). A 10 μL or 20 μl reaction was set up using 50 ng cDNA with 1x TaqMan Fast Advanced Master Mix (Thermo Fisher Scientific, Cat# 4444964) and 1x TaqMan gene expression assay probes. All samples were run in a 384-well plate with three technical replicates per biological replicate. Gene expression was determined using the ΔCt method. Expression levels were calculated using the 2^-ΔCt^ method with TBP (VIC-labeled) as the housekeeping control. P-values were calculated based on 2^-ΔCt^ values from four biological replicates each with three technical replicates using Welch’s-corrected unpaired, two-tailed t-test for comparing WT control to HT variant samples.

### Allele-specific expression of CDKN2A/p16 and CDKN2B in patient-derived cell lines

Haplotype phasing from PacBio WGS was used to phase familial variants with alleles of coding SNPs located in *CDKN2A* and *CDKN2B* transcripts. For patient fibroblasts heterozygous for rs11515 (located in the 3’UTR of *CDKN2A*) and/or rs3217992 (located at the 3’UTR of *CDKN2B*), we phased alleles of these SNPs with chr9 g.22008768C>G (pedigree M) and chr9 g:22008764C>T (pedigree X, **Supplementary Figure 8**), respectively. For assessing ASE in the p16 transcript of *CDKN2A*, no single TaqMan genotyping assay was able to amplify both cDNA and gDNA over rs11515; thus, we employed a Sanger sequencing-based approach. For p16 ASE, genomic DNA was extracted and cDNA was synthesized from total RNA. Both were PCR-amplified over the rs11515 region and subjected to Sanger sequencing.

For RNA isolation, cells from cell culture dishes or from cryo-preserved vials were washed twice with cold Dulbecco’s 1x Phosphate Buffered Saline (PBS; KD Medical, Cat# RGE-3090) on ice and lysed with Trizol LS Reagent (Life Technologies, Cat# 10296010). Trizol was heated to 65°C for 5 min to maximize melanin removal. Following heating, chloroform was added, vortexed, and centrifuged. The aqueous phase was extracted and mixed with an equal volume of 70% EtOH. Samples were then processed using an RNeasy Plus Mini Kit (Qiagen, Cat# 74136) with on-column DNase I digestion per the manufacturer’s protocol (Qiagen, Cat# 79256), and the concentration of isolated RNA was measured via a NanoDrop 8000 spectrophotometer (Thermo Fisher Scientific, Cat#ND-8000-GL). For genomic DNA isolation, the Quick-DNA Miniprep Plus Kit (Zymo Research, Cat# D4069) was used according to the manufacturer’s protocol for Biological Fluids and Cells. DNA concentration and purity were assessed using a Nanodrop 8000 spectrophotometer or Invitrogen Qubit 3 Fluorometer with Qubit dsDNA HS Assay Kit (Thermo Fisher Scientific, Cat #Q32854). Isolated DNA was diluted to 10 ng/µL using nuclease-free ultra-pure water (KD Medical, Cat #RGF-3410) for Sanger sequencing assays.

For Sanger-sequencing based assessment of allele specific expression (p16), complementary DNA (cDNA) was synthesized from 1 μg total RNA in a 20 μl reaction using SuperScript VILO Master Mix Kit (Thermo Fisher Scientific, Cat# 11756050) with a *CDKN2A*-enriching primer (0.5 μΜ, Integrated DNA Technologies; **Supplementary Table 5**). PCR was used to amplify the p16 isoform from 50 ng of cDNA using GoTaq Colorless Master Mix (Promega, Cat#M7822) with 1Μ Βetaine (Thermo Fisher Scientific, Cat# J77507.UCR), and primers (0.5 μΜ, Integrated DNA Technologies; **Supplementary Table 5**). Touchdown PCR was performed under the following conditions: 95°C for 2 min, [95°C for 30 s, 65°C for 30 s, 72°C for 1.5 min] × 10 cycles, [95°C for 30 s, 55°C for 30 s, 72°C for 1.5 min] × 35 cycles, 72°C for 5 min. gDNA over the *CDKN2A* region which covers both transcripts was PCR amplified from 10 ng or 20 ng of gDNA using AmpliTaq Gold 360 Master Mix (Thermo Fisher Scientific, Cat#43-988-81) with 0.5 μΜ primers (Integrated DNA Technologies; **Supplementary Table 5**). Thermocycling conditions were as follows: 95°C for 10 min, [(95°C for 30 s, 60°C for 30 s, 72°C for 1 min]) × 35 cycles, 72°C for 10 min. PCR amplicons from cDNA and gDNA were purified using Mag-Bind Total Pure NGS AMPure beads (Omega Bio-TEK, Cat# M1378-00) followed by Sanger sequencing using the BigDye Terminator v3.1 Cycle Sequencing Kit (Thermo Fisher Scientific, Cat# 4336911) with subsequent purification using Mag-Bind Seq DTR CleanSeq beads (Omega Bio-TEK, Cat# M1300-00S) according to the manufacturer’s instructions. Samples were run on a 3500xl Genetic Analyzer (Thermo Fisher Scientific), and Sanger sequencing chromatograms were visualized using SnapGene software (v4.1.9). p16-specific ASE was determined by obtaining proportion of the peak height of variant haplotype at rs11515 position in both cDNA and DNA. Four biological replicates with one technical replicate for each patient fibroblast cDNA sample were analyzed and compared with five biological replicates of gDNA samples from patient cells. Statistical analysis between cDNA and gDNA samples of patient cells were performed using a Welch-corrected unpaired two-tailed t-test.

### CRISPR-interference (CRISPRi) validation of gene regulation via CTCF binding motif

To validate gene regulatory functions via the CTCF binding motif located around the family M variant, we performed CRISPR interference (CRISPRi) assays in C283T cells stably expressing dCas9-KRAB and in human primary melanocytes (C87) via transient expression of dCas9-KRAB. Briefly, C283T cells were infected with a lentiviral vector pLX_311-KRAB-dCas9 (gift from John Doench, William Hahn, and David Root; Addgene plasmid #96918; RRID: Addgene_96918) followed by monoclonal cell selection. Stable expression and functional activity of dCas9-KRAB was confirmed in the C283T clone used for CRISPRi experiments. For C87 cells, lentiviral particles expressing pLX-311-KRAB-dCas9 was co-infected with lentivirus expressing sgRNAs and the expression of dCas9-KRAB was confirmed by Western Blot.

Two different sgRNAs were designed to target the CTCF binding motif and are listed in **Supplementary Table 5** and shown in **Supplementary Figure 10**. Lentivirus particles were made by co-transfecting and expressing both sgRNAs targeting the CTCF binding motif. Additionally, two non-targeting sgRNAs (NTC1, NTC2) were used as controls and are listed in **Supplementary Table 5.** sgRNAs were ligated into the lentiviral vector pXPR-050 (gift from John Doench and David Root, Addgene plasmid #96925; RRID: Addgene_96925). Cells were infected with either lentiviral particles encoding both sgRNAs targeting CTCF or mixed viral particles expressing NTC1 or NTC2 sgRNA, and at 24 h, 1.2 μg/mL of puromycin (Thermo Fisher Scientific, Cat# A1113803) was added for selection. After two days of antibiotic selection, cells were collected for RNA isolation. Total RNA was isolated using an RNeasy Plus Mini Kit (Qiagen, Cat#74136) and cDNA was generated with SuperScript IV VILO Master Mix (Thermo Fisher Scientific, Cat#11756050). Three infections were performed for either sgRNAs targeting CTCF or NTC controls. mRNA levels of p14, p16, *CDKN2B*, *MTAP*, *CDKN2B-AS1*, *and DMRTA1*, were measured by TaqMan assay (Thermo Scientific) and normalized to *TBP* levels. 2^-ΔΔCt^ were calculated to determine relative gene expression levels between NTC controls and sgRNAs targeting CTCF. Each biological replicate comprised of averaged qPCR triplicates (technical replicates) 2^-ΔCt^ values were used for statistical testing using an unpaired two-tailed t-test.

### UK Biobank analysis

UK Biobank (UKBB) is a large population-based retrospective and prospective study of participants recruited from age 40-69 years; 469,376 participants were exome-sequenced with health record data. Human subjects’ protection and review was through the North West Multi-centre Research Ethics Committee [93, 94]. Thirteen *CDKN2B* germline variants were extracted from field 23157 (population level exome OQFE variants, pVCF format – final exome release) and filtered for variants with a genotype quality (GQ) > 30 and an allelic balance (AB) Het >0.3 or < 0.7. Variants were confirmed using the Integrated Genomics Viewer [95].

To select individuals who were cancer-free from the full exome cohort, participants with a “C” and/or D03 code in ICD10 or 140-208 in ICD9 from cancer registry (field 40006, 40013), death registry (field 40001), and EHR (field 41270, 41271) were excluded. To select individuals with melanoma, participants with ICD10 C43, D03, and/or ICD9 172 codes were included. In total, nine of the 13 variants in this region were found in the case-control set (**Table 1**). For statistical analysis, a Fisher’s exact test was used in a 2 x 2 contingency table as well as a general linear model using age and sex as covariates as implemented in the R glm function.

## Data availability

WES and targeted sequencing data from melanoma families and targeted sequencing data from melanoma cases from the Prostate, Lung, Colorectal and Ovarian Cancer Screening [47] trial that were previously published [46, 47] are available via the NCBI database of Genotypes and Phenotypes (dbGAP; phs001177.v2.p1). RNA-seq, ChiP-seq, and PacBio longreads sequencing data from subjects in Pedigree M are available in dbGAP (phs001177.v3).

RNA sequencing data from human primary melanocytes [55] is available via dbGAP (dbGAP phs001500.v1). RNA sequencing data from melanoma cell lines [56] is available from NCBI Gene Expression Omnibus (GEO; GSE140673). RNA Sequencing data for The Cancer Genome Atlas skin cutaneous melanoma (SKCM) project [30] is available via the National Cancer Institute Genomic Data Commons Portal (https://portal.gdc.cancer.gov/projects/TCGA-SKCM).

Capture Hi-C data generated from primary melanocytes [64] is available through gEMBL-EBI ArrayExpress (E-MTAB-15079).

UK Biobank (UKBB) data is available through the UKBB Research Analysis Platform (https://www.ukbiobank.ac.uk/use-our-data/research-analysis-platform/).

## Acknowledgements

This research was supported in part by the Intramural Research Program of the National Institutes of Health (NIH). The contributions of the NIH author(s) are considered Works of the United States Government. This project has been funded in part with Federal funds from the National Cancer Institute, National Institutes of Health, under Contract No. 75N91019D00024. The findings and conclusions of this publication do not necessarily reflect the views or policies of the Department of Health and Human Services, nor does mention of trade names, commercial products, or organizations imply endorsement by the U.S. Government. This work utilized the Biowulf cluster computing system at the NIH. The results appearing here are in part based on data generated by the TCGA Research Network. We would like to thank members at the National Cancer Institute Cancer Genomics Research Laboratory (CGR) for their help with sequencing efforts. This research has been conducted using the UK Biobank Resource under Application Number 54389. The authors express sincere appreciation to all Cancer Prevention Study-II participants, and to each member of the study and biospecimen management group. The authors would like to acknowledge the contribution to this study from central cancer registries supported through the Centers for Disease Control and Prevention’s National Program of Cancer Registries and cancer registries supported by the National Cancer Institute’s Surveillance Epidemiology and End Results Program.

---

**Supplementary Figure 1.**
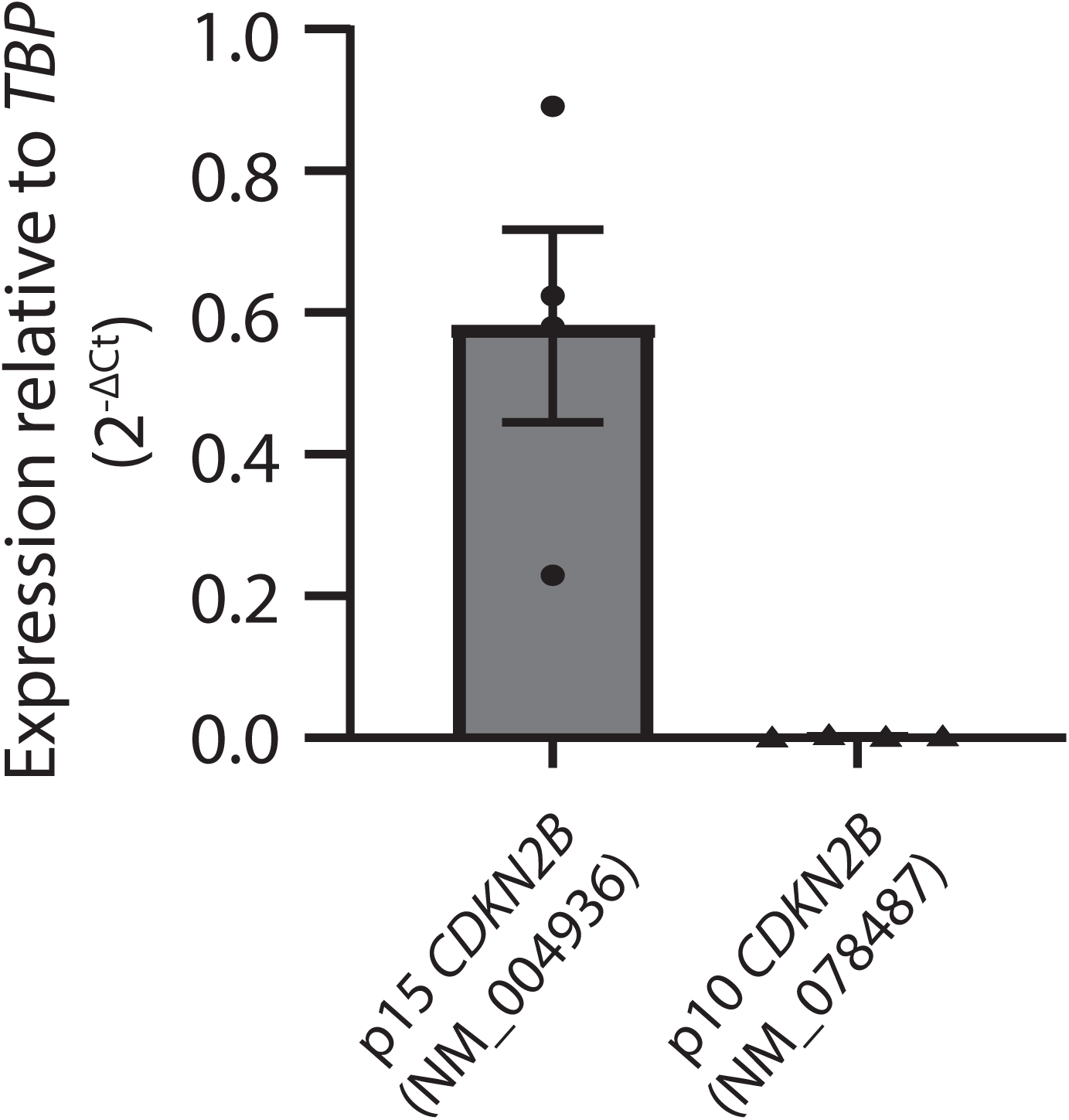
Expression of *CDKN2B* p15 and p10 isoforms in immortalized human melanocytes. *CDKN2B* transcript expression was assessed by isoform-specific TaqMan qRT-PCR assays normalized relative to expression of TBP. Each data point shows the average of three technical replicates for four biological replicates.

**Supplementary Figure 2.**
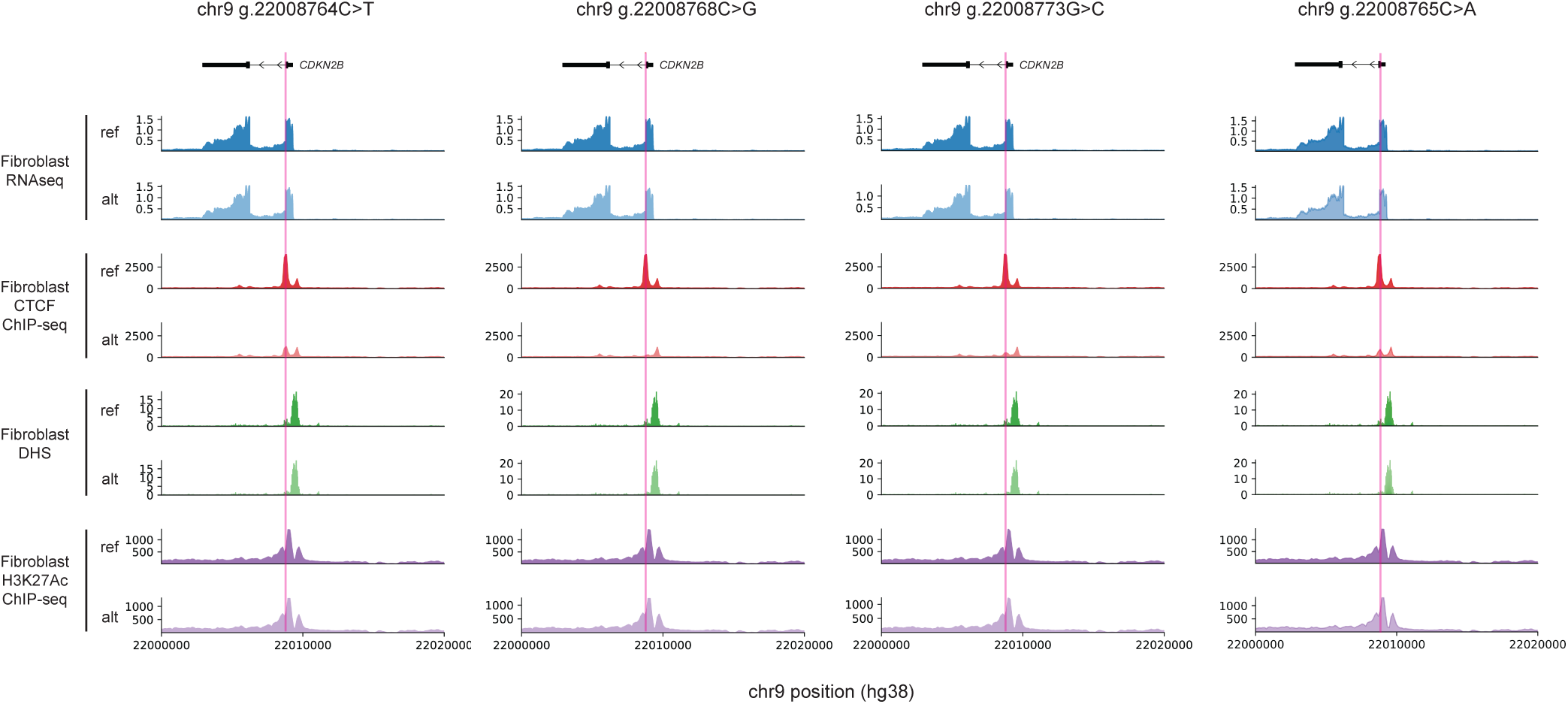
*CDKN2B* variants are predicted to alter CTCF binding and chromatin accessibility. AlphaGenome predictions of the consequences of each of the variants identified in American melanoma cases and UKBB on transcription (RNA-seq), CTCF-binding (CTCF ChIP-seq), chromatin accessibility as assessed by DNAseI hypersensitivity sequencing (DHS), and histone mark H3K27Ac (H3K27Ac ChIP-seq). The predictions suggest predicted loss of CTCF binding as well as a modest reduction of chromatin accessibility over each of the variant positions for alternative alleles (alt) relative to the reference (ref). Variant position is highlighted in pink; genes in black at the top of each panel.

**Supplementary Figure 3.**
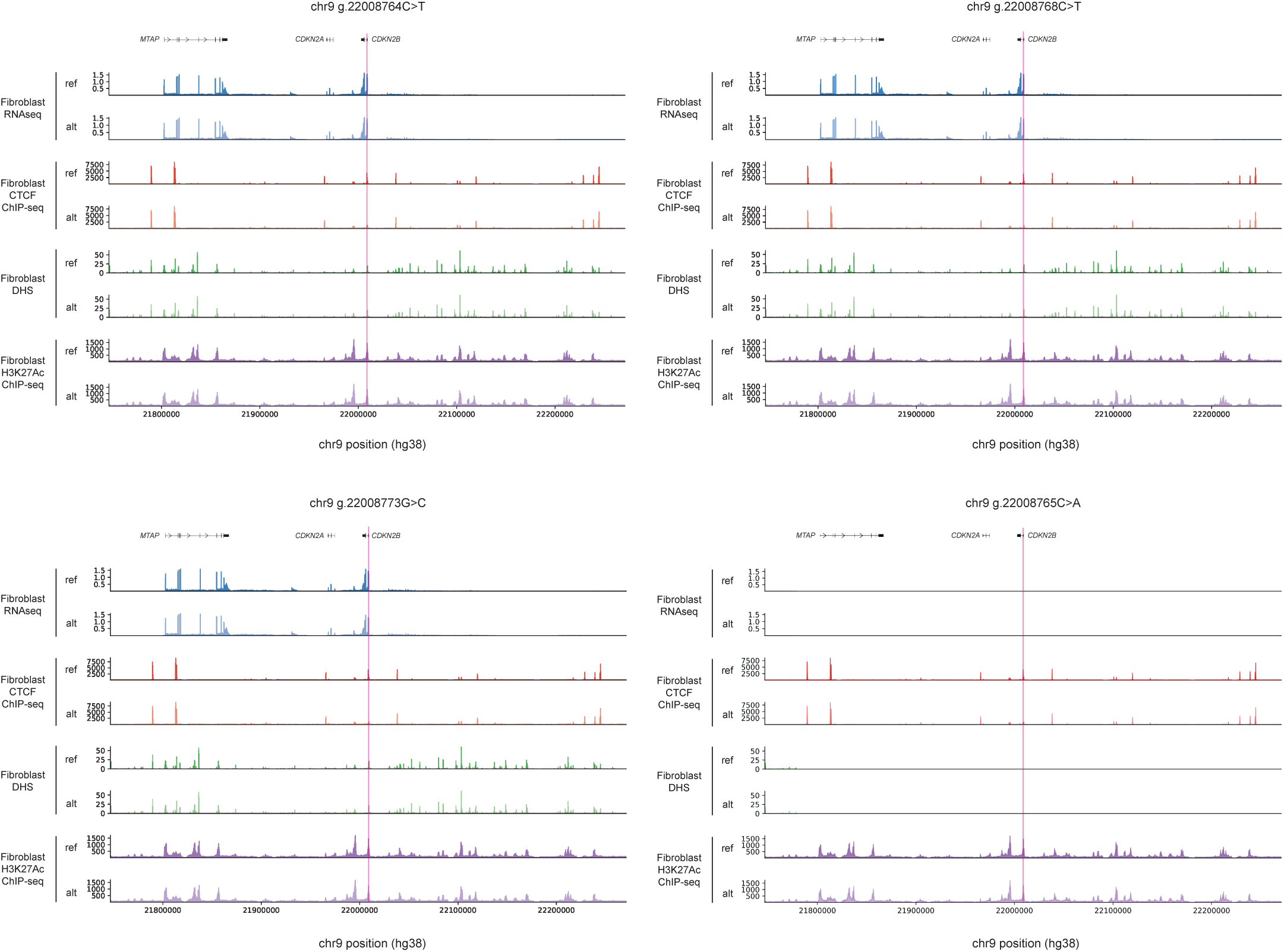
*CDKN2B* variants are predicted to alter CTCF binding and chromatin accessibility. A zoomed-out view of AlphaGenome predictions of the consequences of each of the variants identified in American melanoma cases and UKBB on transcription (RNA-seq), CTCF-binding (CTCF ChIP-seq), chromatin accessibility as assessed by DNAseI hypersensitivity sequencing (DHS), and histone mark H3K27Ac (H3K27Ac ChIP-seq). The predictions suggest predicted loss of CTCF binding as well as a modest reduction of chromatin accessibility over each of the variant positions for alternative alleles (alt) relative to the reference (ref). Variant position is highlighted in pink; genes in black at the top of each panel.

**Supplementary Figure 4.**
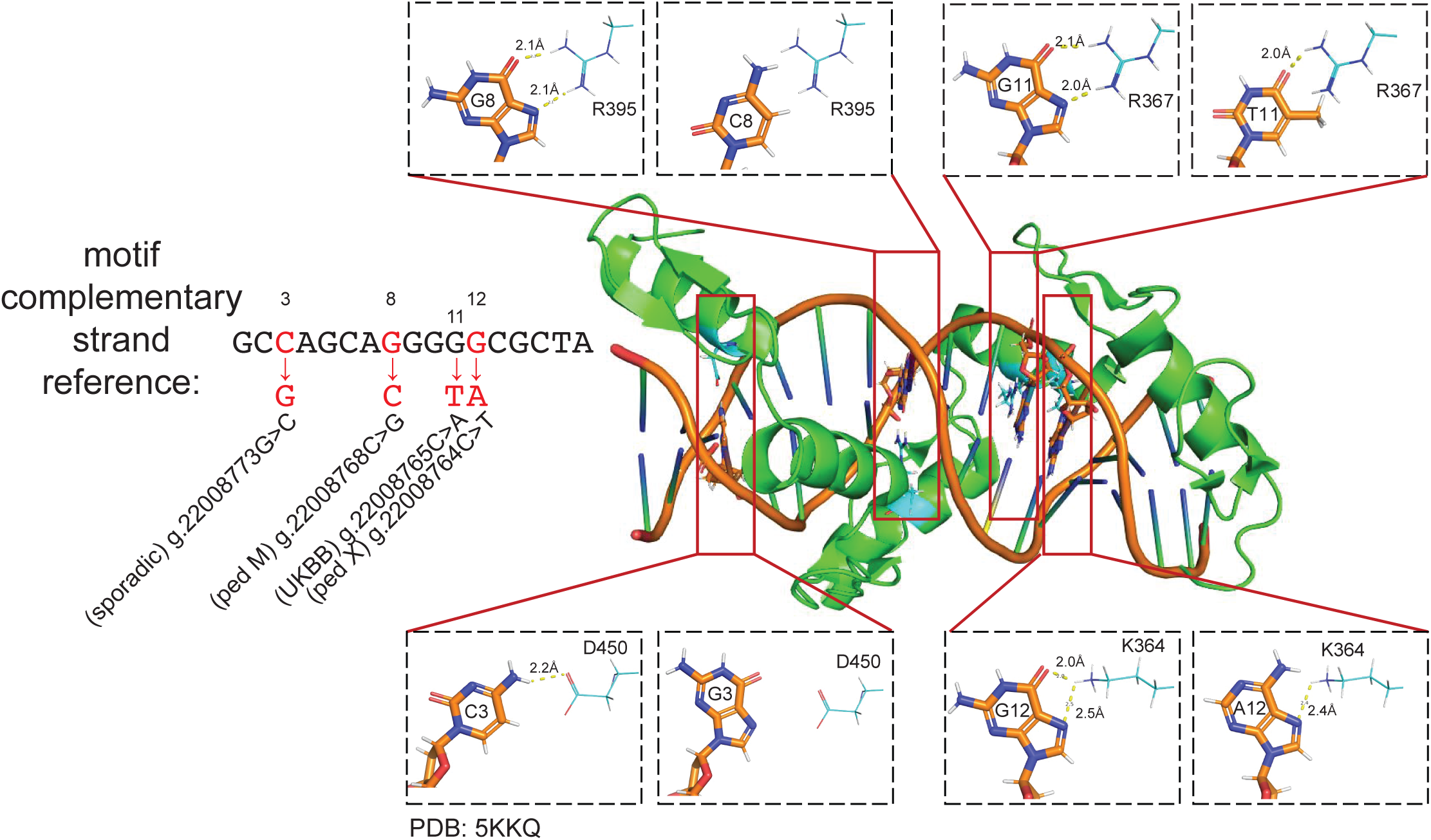
Melanoma risk variants alter CTCF interaction with DNA. Cytosine at position 3 (g.22008773G), guanine at position 8 (g.22008768C), guanine at position 11 (g22008765C), and guanine at position 12 (g.22008764C) in the complementary strand of the CTCF DNA-binding motif form hydrogen bonds (yellow dotted line) with D450, R395, R367, and K364 of the CTCF protein, respectively. These are lost upon their respective base substitutions observed in melanoma cases. Structures are generated from PDB:5KKQ. The minus-strand of the CTCF-binding site is shown and labeled with respective base substitutions.

**Supplementary Figure 5.**
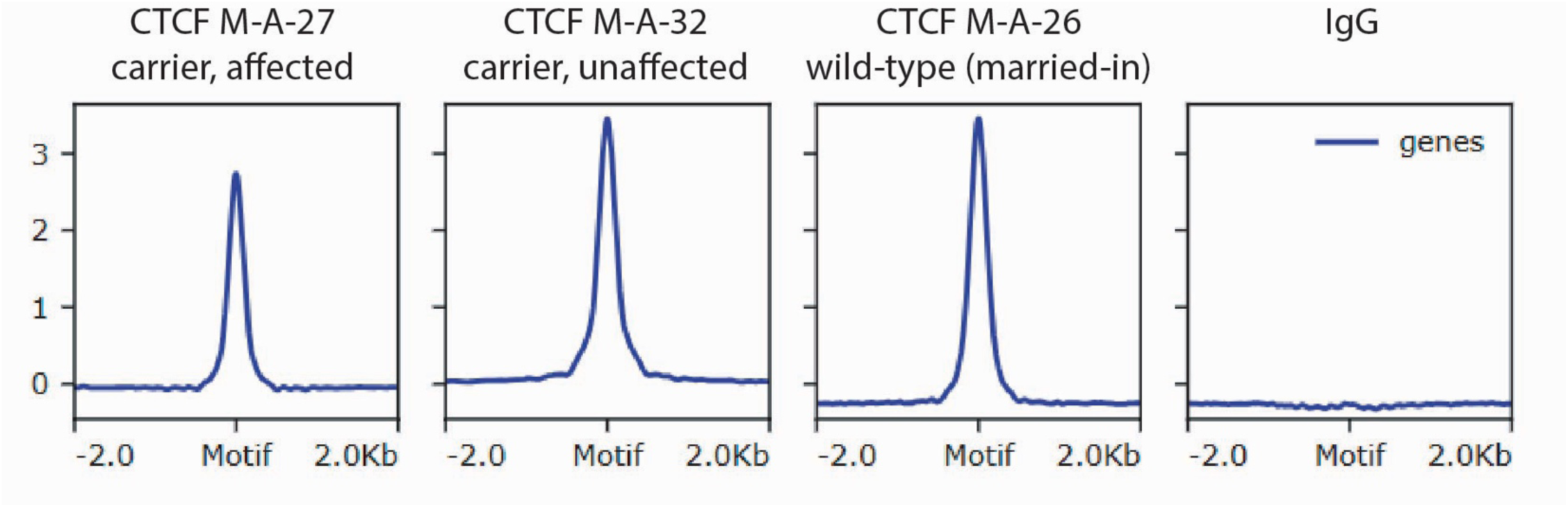
CTCF ChIP seq assessment of fibroblasts from carriers of familial sequence variants. Peak enrichment over known CTCF binding sites is shown by CTCF ChIP-seq experiments from fibroblasts from two carriers (M-A-27 and M-A-37) and one non-carrier (M-A-26).

**Supplementary Figure 6.**
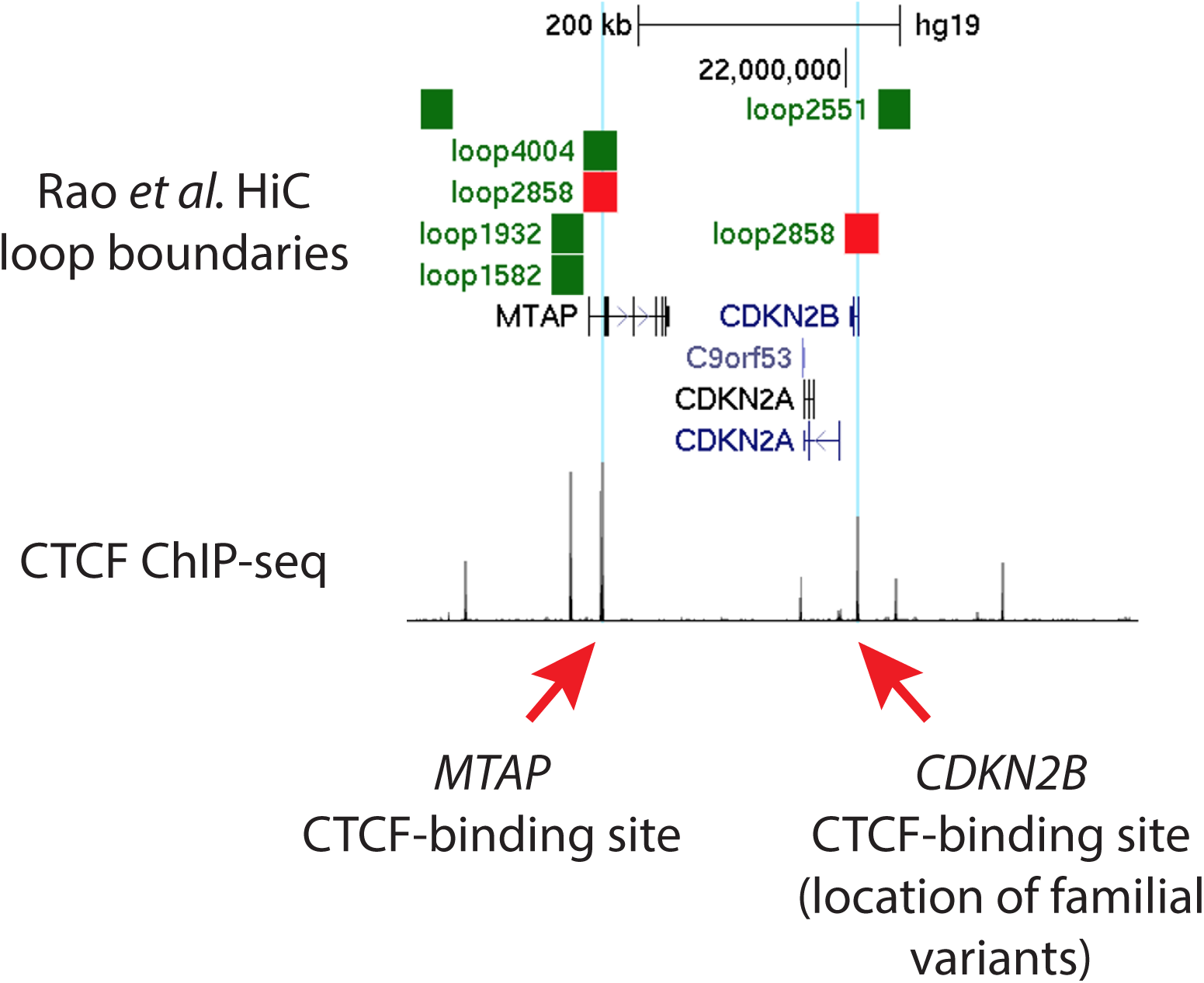
UCSC Genome Browser representation of CTCF ChIP coverage as signal normalized to input and HiC loops identified by Rao and colleagues (upper). The Rao loop that covers the *CDKN2B* CTCF binding site (ID loop 2858) is highlighted in red. The *CDKN2B* CTCF binding site that contains mutations is highlighted in blue and indicated with a red arrow. The MTAP CTCF binding site that forms a loop with *CDKN2B* one is also highlighted in blue and indicated with a red arrow. Scale bar indicating length equivalent to 200kb is given at the top.

**Supplementary Figure 7.**
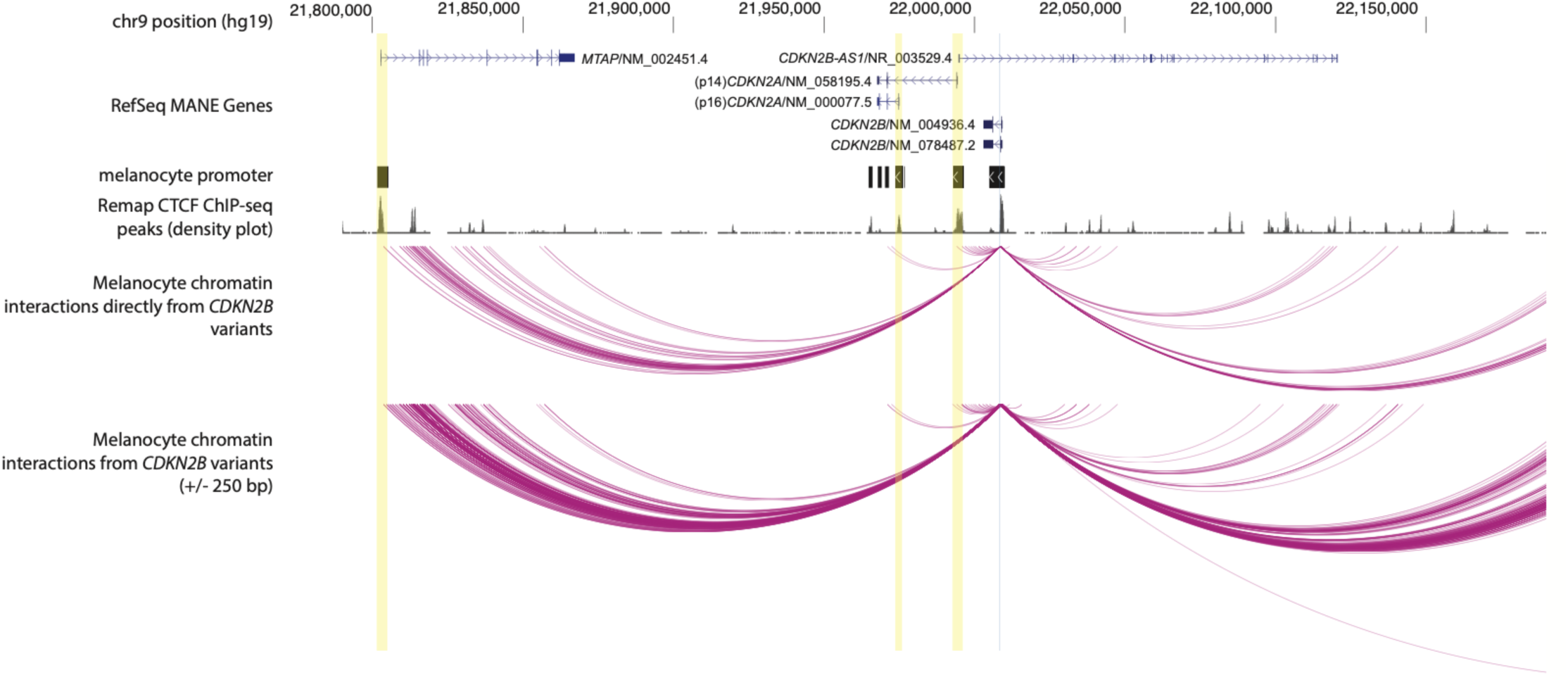
Chromatin interactions with the *CDKN2B* variant region. Chromatin interactions from the region harboring melanoma risk-associated *CDKN2B* variants in human primary melanocytes. Data are from a previously published capture-HiC assay from Thakur and colleagues. Interactions were assessed using capture-baits tiled across the 9p21 region. Data are filtered to show only the most highly-scored (80^th^ percentile or higher of significant interactions) interactions directly from the region harboring the variants (upper panel, highlighted in light blue) or from a larger 500 bp window surrounding the variants (lower panel). Annotated melanocyte promoter regions (ChromHMM) from the Roadmap Epigenome Project are shown, as are the density of annotated ENCODE CTCF-binding sites from ReMap. Promoters of the p16 and p14 transcripts of *CDKN2A*, as well as the promoter of MTAP, are highlighted in yellow. Coordinates are hg38.

**Supplementary Figure 8.**
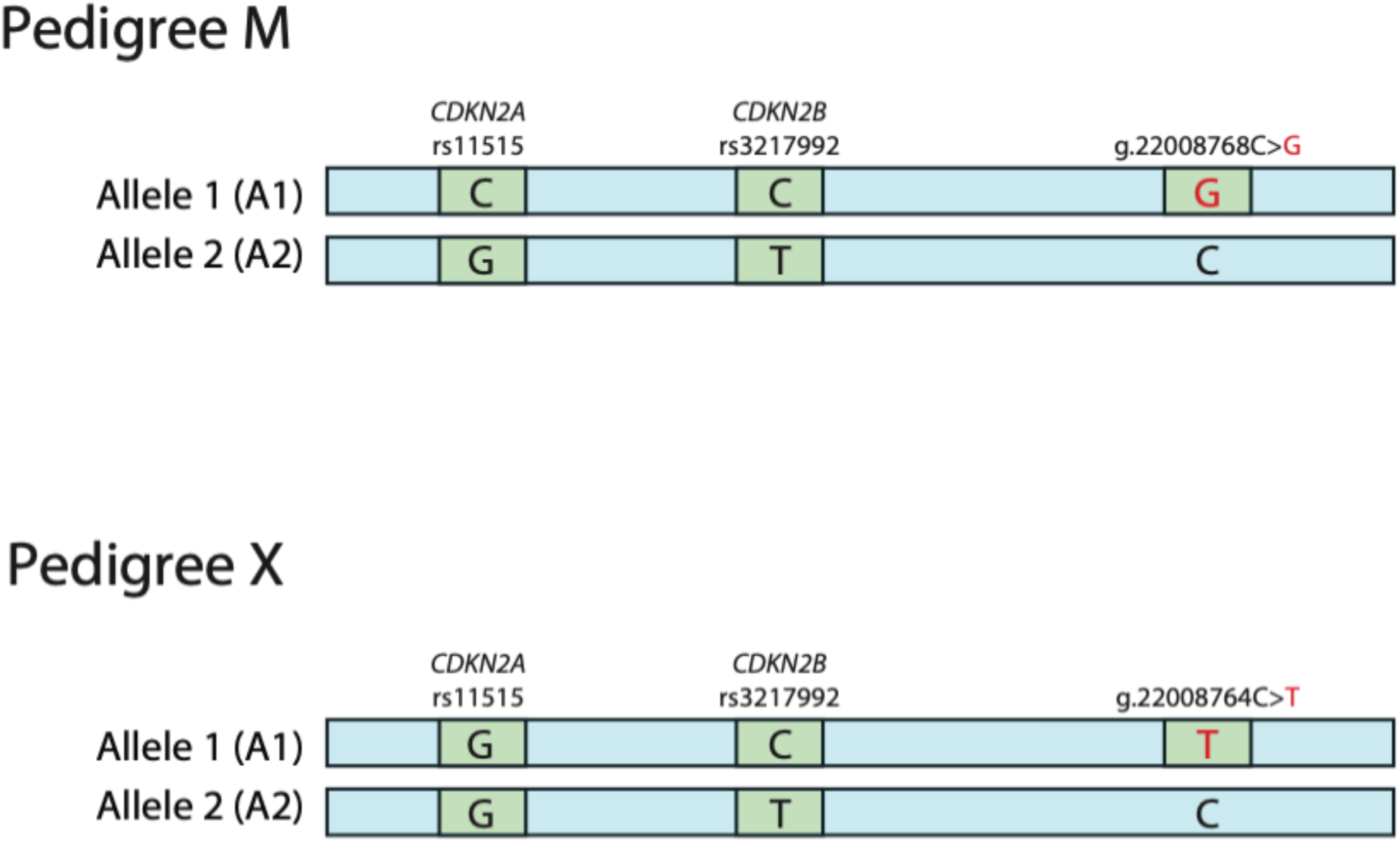
Schematic of SNP allele phasing relative to familial *CDKN2B* variants. Phase of variants for patient-derived samples from melanoma families was assessed via long-read whole-genome sequencing (see **Methods**).

**Supplementary Figure 9.**
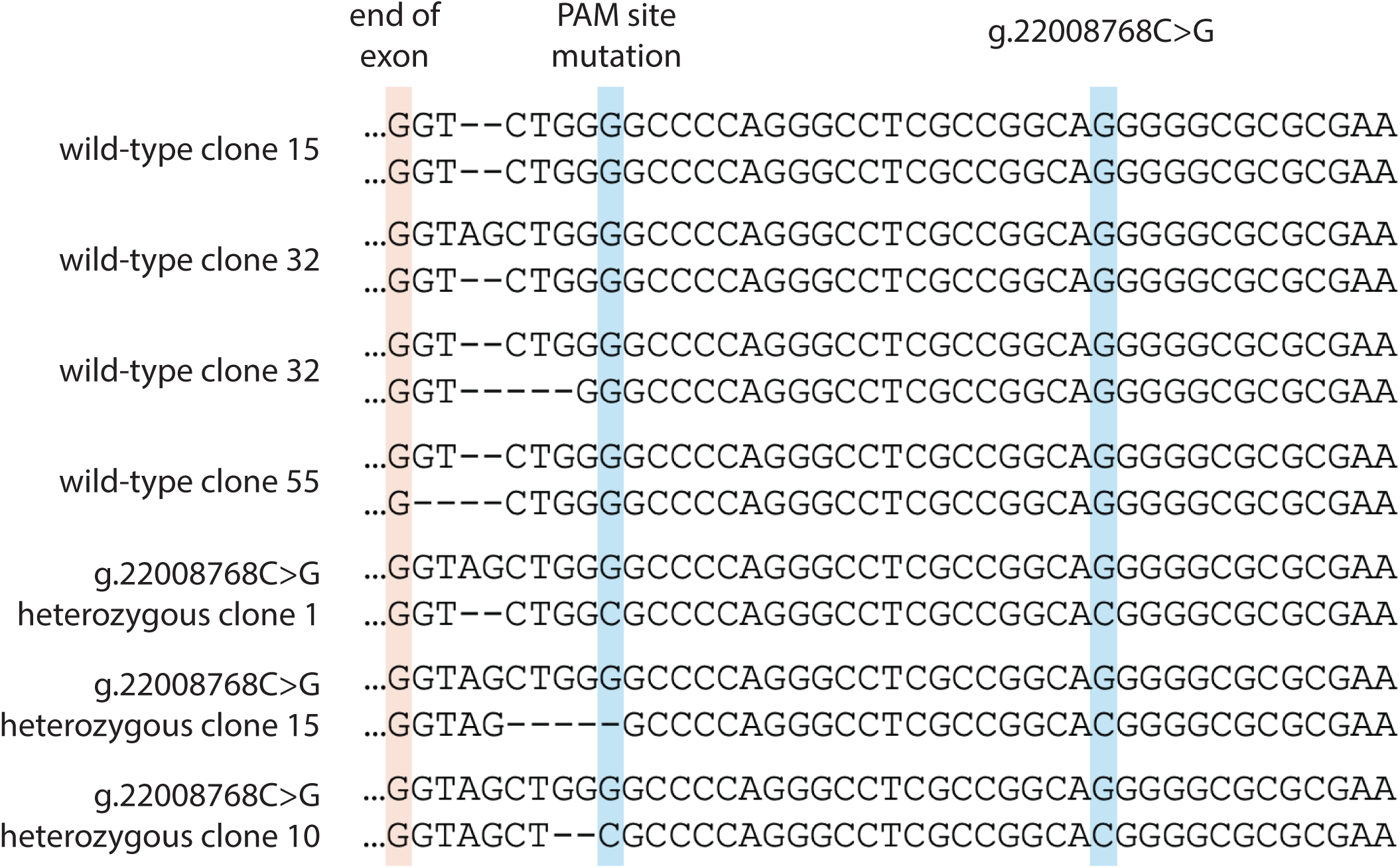
DNA sequence deconvolution for CRISPR-Cas9-edited primary melanocytes. Each of two alleles for each clone as assessed via Sanger sequencing. Sequence is shown relative to the direction of the *CDKN2B* transcript (minus strand of the genome); a successful edit for the g.22008768C>G variant is shown as a “C” allele. The relative sites of the end of the first exon of the CDKN2B p15 transcript and engineered PAM site mutation are shown. Deleted based are denoted by “-“.

**Supplementary Figure 10.**
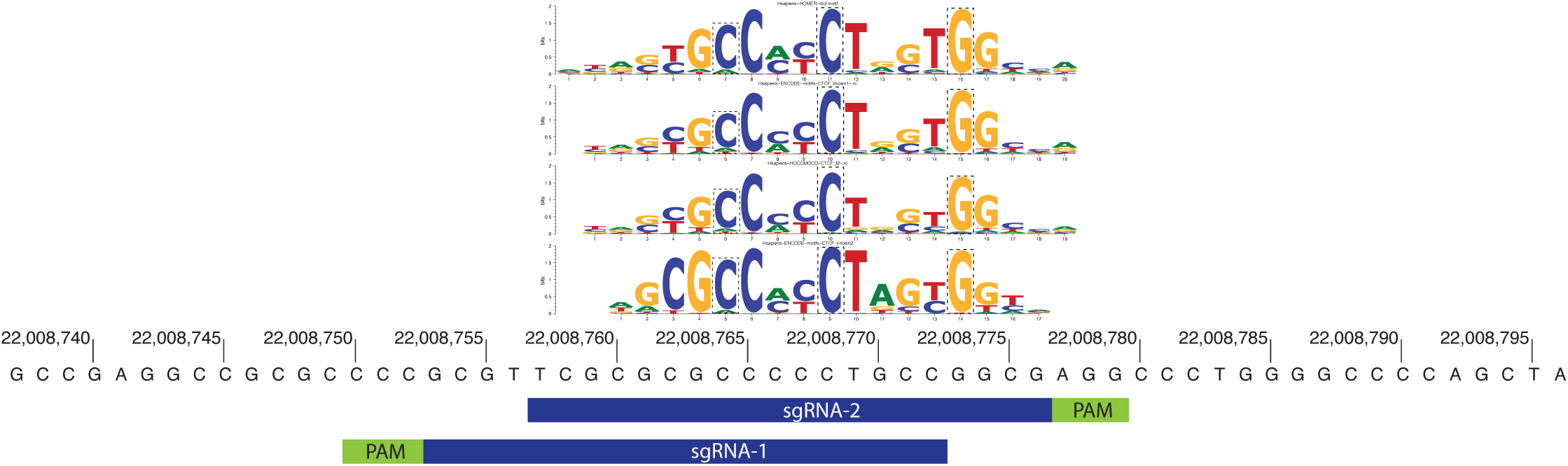
Location of small guide RNAs (sgRNAs) used for CRISPR-inhibition experiments. Locations of guides are shown relative to the CTCF motif. Locations of the three risk-associated variants is denoted on the motif by based outlined in a dashed rectangle.

## References

1. Garber, J.E. and K. Offit, Hereditary cancer predisposition syndromes. J Clin Oncol, 2005. 23(2): p. 276–92.

2. Shekar, S.N., D.L. Duffy, P. Youl, A.J. Baxter, M. Kvaskoff, D.C. Whiteman, A.C. Green, M.C. Hughes, N.K. Hayward, M. Coates, and N.G. Martin, A population-based study of Australian twins with melanoma suggests a strong genetic contribution to liability. J Invest Dermatol, 2009. 129(9): p. 2211–9.

3. Mucci, L.A., J.B. Hjelmborg, J.R. Harris, K. Czene, D.J. Havelick, T. Scheike, R.E. Graff, Holst, S. Moller, R.H. Unger, C. McIntosh, E. Nuttall, I. Brandt, K.L. Penney, M. Hartman, P. Kraft, G. Parmigiani, K. Christensen, M. Koskenvuo, N.V. Holm, K. Heikkila, E. Pukkala, A. Skytthe, H.O. Adami, J. Kaprio, and C. Nordic Twin Study of Cancer, Familial Risk and Heritability of Cancer Among Twins in Nordic Countries. JAMA, 2016. 315(1): p. 68–76.

4. Cannon-Albright, L.A., D.E. Goldgar, L.J. Meyer, C.M. Lewis, D.E. Anderson, J.W. Fountain, M.E. Hegi, R.W. Wiseman, E.M. Petty, A.E. Bale, and, et al., Assignment of a locus for familial melanoma, MLM, to chromosome 9p13-p22. Science, 1992. 258(5085): p. 1148–52.

5. Hussussian, C.J., J.P. Struewing, A.M. Goldstein, P.A. Higgins, D.S. Ally, M.D. Sheahan, W.H. Clark, Jr., M.A. Tucker, and N.C. Dracopoli, Germline p16 mutations in familial melanoma. Nat Genet, 1994. 8(1): p. 15–21.

6. Ranade, K., C.J. Hussussian, R.S. Sikorski, H.E. Varmus, A.M. Goldstein, M.A. Tucker, M. Serrano, G.J. Hannon, D. Beach, and N.C. Dracopoli, Mutations associated with familial melanoma impair p16INK4 function. Nat Genet, 1995. 10(1): p. 114–6.

7. Goldstein, A.M. and M.A. Tucker, Genetic epidemiology of cutaneous melanoma: a global perspective. Arch Dermatol, 2001. 137(11): p. 1493–6.

8. Kefford, R.F., J.A. Newton Bishop, W. Bergman, and M.A. Tucker, Counseling and DNA testing for individuals perceived to be genetically predisposed to melanoma: A consensus statement of the Melanoma Genetics Consortium. J Clin Oncol, 1999. 17(10): p. 3245–51.

9. Goldstein, A.M., M. Chan, M. Harland, N.K. Hayward, F. Demenais, D.T. Bishop, E. Azizi, W. Bergman, G. Bianchi-Scarra, W. Bruno, D. Calista, L.A. Albright, V. Chaudru, A. Chompret, F. Cuellar, D.E. Elder, P. Ghiorzo, E.M. Gillanders, N.A. Gruis, J. Hansson, D. Hogg, E.A. Holland, P.A. Kanetsky, R.F. Kefford, M.T. Landi, J. Lang, S.A. Leachman, R.M. MacKie, V. Magnusson, G.J. Mann, J.N. Bishop, J.M. Palmer, S. Puig, J.A. Puig-Butille, M. Stark, H. Tsao, M.A. Tucker, L. Whitaker, E. Yakobson, G. Lund Melanoma Study, and C. Melanoma Genetics, Features associated with germline CDKN2A mutations: a GenoMEL study of melanoma-prone families from three continents. J Med Genet, 2007. 44(2): p. 99–106.

10. Goldstein, A.M. and M.A. Tucker, Screening for CDKN2A mutations in hereditary melanoma. J Natl Cancer Inst, 1997. 89(10): p. 676–8.

11. Sargen, M.R., H. Helgadottir, X.R. Yang, M. Harland, J.N. Hatton, K. Jones, B.D. Hicks, A. Hutchinson, M. Curry, M.A. Tucker, A.M. Goldstein, and R.M. Pfeiffer, Impact of Transcript (p16/p14ARF) Alteration on Cancer Risk in CDKN2A Germline Pathogenic Variant Carriers. JNCI Cancer Spectr, 2022. 6(6).

12. Goldstein, A.M., M. Chan, M. Harland, E.M. Gillanders, N.K. Hayward, M.F. Avril, E. Azizi, G. Bianchi-Scarra, D.T. Bishop, B. Bressac-de Paillerets, W. Bruno, D. Calista, L.A. Cannon Albright, F. Demenais, D.E. Elder, P. Ghiorzo, N.A. Gruis, J. Hansson, D. Hogg, E.A. Holland, P.A. Kanetsky, R.F. Kefford, M.T. Landi, J. Lang, S.A. Leachman, R.M. Mackie, V. Magnusson, G.J. Mann, K. Niendorf, J. Newton Bishop, J.M. Palmer, S. Puig, J.A. Puig-Butille, F.A. de Snoo, M. Stark, H. Tsao, M.A. Tucker, L. Whitaker, E. Yakobson, and C. Melanoma Genetics, High-risk melanoma susceptibility genes and pancreatic cancer, neural system tumors, and uveal melanoma across GenoMEL. Cancer Res, 2006. 66(20): p. 9818–28.

13. Molven, A., M.B. Grimstvedt, S.J. Steine, M. Harland, M.F. Avril, N.K. Hayward, and L.A. Akslen, A large Norwegian family with inherited malignant melanoma, multiple atypical nevi, and CDK4 mutation. Genes Chromosomes Cancer, 2005. 44(1): p. 10–8.

14. Soufir, N., M.F. Avril, A. Chompret, F. Demenais, J. Bombled, A. Spatz, D. Stoppa-Lyonnet, J. Benard, and B. Bressac-de Paillerets, Prevalence of p16 and CDK4 germline mutations in 48 melanoma-prone families in France. The French Familial Melanoma Study Group. Hum Mol Genet, 1998. 7(2): p. 209–16.

15. Zuo, L., J. Weger, Q. Yang, A.M. Goldstein, M.A. Tucker, G.J. Walker, N. Hayward, and N.C. Dracopoli, Germline mutations in the p16INK4a binding domain of CDK4 in familial melanoma. Nat Genet, 1996. 12(1): p. 97–9.

16. Goldstein, A.M., A. Chidambaram, A. Halpern, E.A. Holly, I.D. Guerry, R. Sagebiel, D.E. Elder, and M.A. Tucker, Rarity of CDK4 germline mutations in familial melanoma. Melanoma Res, 2002. 12(1): p. 51–5.

17. Horn, S., A. Figl, P.S. Rachakonda, C. Fischer, A. Sucker, A. Gast, S. Kadel, I. Moll, E. Nagore, K. Hemminki, D. Schadendorf, and R. Kumar, TERT promoter mutations in familial and sporadic melanoma. Science, 2013. 339(6122): p. 959–61.

18. Harland, M., M. Petljak, C.D. Robles-Espinoza, Z. Ding, N.A. Gruis, R. van Doorn, K.A. Pooley, A.M. Dunning, L.G. Aoude, K.A. Wadt, A.M. Gerdes, K.M. Brown, N.K. Hayward, J.A. Newton-Bishop, D.J. Adams, and D.T. Bishop, Germline TERT promoter mutations are rare in familial melanoma. Fam Cancer, 2016. 15(1): p. 139–44.

19. Shi, J., X.R. Yang, B. Ballew, M. Rotunno, D. Calista, M.C. Fargnoli, P. Ghiorzo, B. Bressac-de Paillerets, E. Nagore, M.F. Avril, N.E. Caporaso, M.L. McMaster, M. Cullen, Z. Wang, X. Zhang, N.D.C.S.W. Group, N.D.C.G.R. Laboratory, G. French Familial Melanoma Study, W. Bruno, L. Pastorino, P. Queirolo, J. Banuls-Roca, Z. Garcia-Casado, A. Vaysse, H. Mohamdi, Y. Riazalhosseini, M. Foglio, F. Jouenne, X. Hua, P.L. Hyland, J. Yin, H. Vallabhaneni, W. Chai, P. Minghetti, C. Pellegrini, S. Ravichandran, A. Eggermont, M. Lathrop, K. Peris, G.B. Scarra, G. Landi, S.A. Savage, J.N. Sampson, J. He, M. Yeager, L.R. Goldin, F. Demenais, S.J. Chanock, M.A. Tucker, A.M. Goldstein, Y. Liu, and M.T. Landi, Rare missense variants in POT1 predispose to familial cutaneous malignant melanoma. Nat Genet, 2014. 46(5): p. 482–6.

20. Robles-Espinoza, C.D., M. Harland, A.J. Ramsay, L.G. Aoude, V. Quesada, Z. Ding, K.A., Pooley, A.L. Pritchard, J.C. Tiffen, M. Petljak, J.M. Palmer, J. Symmons, P. Johansson, M.S. Stark, M.G. Gartside, H. Snowden, G.W. Montgomery, N.G. Martin, J.Z. Liu, J. Choi, M. Makowski, K.M. Brown, A.M. Dunning, T.M. Keane, C. Lopez-Otin, N.A. Gruis, N.K. Hayward, D.T. Bishop, J.A. Newton-Bishop, and D.J. Adams, POT1 loss-of-function variants predispose to familial melanoma. Nat Genet, 2014. 46(5): p. 478–481.

21. Aoude, L.G., A.L. Pritchard, C.D. Robles-Espinoza, K. Wadt, M. Harland, J. Choi, M. Gartside, V. Quesada, P. Johansson, J.M. Palmer, A.J. Ramsay, X. Zhang, K. Jones, J. Symmons, E.A. Holland, H. Schmid, V. Bonazzi, S. Woods, K. Dutton-Regester, M.S. Stark, H. Snowden, R. van Doorn, G.W. Montgomery, N.G. Martin, T.M. Keane, C. Lopez-Otin, A.M. Gerdes, H. Olsson, C. Ingvar, A. Borg, N.A. Gruis, J.M. Trent, G. Jonsson, D.T. Bishop, G.J. Mann, J.A. Newton-Bishop, K.M. Brown, D.J. Adams, and N.K. Hayward, Nonsense mutations in the shelterin complex genes ACD and TERF2IP in familial melanoma. J Natl Cancer Inst, 2015. 107(2).

22. Jensen, M.R., A.M. Jelsig, A.M. Gerdes, L.R. Holmich, K.H. Kainu, H.F. Lorentzen, M.H. Hansen, M. Bak, P.A. Johansson, N.K. Hayward, T. Van Overeem Hansen, and K.A.W. Wadt, TINF2 is a major susceptibility gene in Danish patients with multiple primary melanoma. HGG Adv, 2023. 4(4): p. 100225.

23. Liu, L., A.M. Goldstein, M.A. Tucker, H. Brill, N.A. Gruis, D. Hogg, and N.J. Lassam, Affected members of melanoma-prone families with linkage to 9p21 but lacking mutations in CDKN2A do not harbor mutations in the coding regions of either CDKN2B or p19ARF. Genes Chromosomes Cancer, 1997. 19(1): p. 52–4.

24. Stone, S., P. Dayananth, P. Jiang, J.M. Weaver-Feldhaus, S.V. Tavtigian, L. Cannon-Albright, and A. Kamb, Genomic structure, expression and mutational analysis of the P15 (MTS2) gene. Oncogene, 1995. 11(5): p. 987–91.

25. Taylor, N.J., N. Mitra, A.M. Goldstein, M.A. Tucker, M.F. Avril, E. Azizi, W. Bergman, D.T. Bishop, B. Bressac-de Paillerets, W. Bruno, D. Calista, L.A. Cannon-Albright, F. Cuellar, A.E. Cust, F. Demenais, D.E. Elder, A.M. Gerdes, P. Ghiorzo, T.C. Grazziotin, J. Hansson, M. Harland, N.K. Hayward, M. Hocevar, V. Hoiom, C. Ingvar, M.T. Landi, G. Landman, A. Larre-Borges, S.A. Leachman, G.J. Mann, E. Nagore, H. Olsson, J.M. Palmer, B. Peric, D. Pjanova, A. Pritchard, S. Puig, N. van der Stoep, K.A.W. Wadt, L. Whitaker, X.R. Yang, J.A. Newton Bishop, N.A. Gruis, P.A. Kanetsky, and M.E.L.S.G. Geno, Germline Variation at CDKN2A and Associations with Nevus Phenotypes among Members of Melanoma Families. J Invest Dermatol, 2017. 137(12): p. 2606–2612.

26. Khurana, E., Y. Fu, D. Chakravarty, F. Demichelis, M.A. Rubin, and M. Gerstein, Role of non-coding sequence variants in cancer. Nat Rev Genet, 2016. 17(2): p. 93–108.

27. Harland, M., S. Mistry, D.T. Bishop, and J.A. Bishop, A deep intronic mutation in CDKN2A is associated with disease in a subset of melanoma pedigrees. Hum Mol Genet, 2001. 10(23): p. 2679–86.

28. Shi, J., W. Zhou, B. Zhu, P.L. Hyland, H. Bennett, Y. Xiao, X. Zhang, L.S. Burke, L. Song, C.H. Hsu, C. Yan, Q. Chen, D. Meerzaman, C.L. Dagnall, L. Burdette, B. Hicks, N.D. Freedman, S.J. Chanock, M. Yeager, M.A. Tucker, A.M. Goldstein, and X.R. Yang, Rare Germline Copy Number Variations and Disease Susceptibility in Familial Melanoma. J Invest Dermatol, 2016. 136(12): p. 2436–2443.

29. Lesueur, F., M. de Lichy, M. Barrois, G. Durand, J. Bombled, M.F. Avril, A. Chompret, F. Boitier, G.M. Lenoir, G. French Familial Melanoma Study, B. Bressac-de Paillerets, M. Baccard, B. Bachollet, P. Berthet, V. Bonadona, J.M. Bonnetblanc, O. Caron, J. Chevrant-Breton, J.F. Cuny, S. Dalle, M. Delaunay, L. Demange, J. De Quatrebarbes, J.F. Dore, M. Frenay, J.P. Fricker, M. Gauthier-Villars, P. Gesta, S. Giraud, P. Gorry, F. Grange, A. Green, L. Huiart, N. Janin, P. Joly, D. Kerob, C. Lasset, D. Leroux, J.M. Limacher, M. Longy, S. Mansard, K. Marrou, T. Martin-Denavit, C. Mateus, E. Maubec, L. Olivier-Faivre, V. Orlandini, P. Pujol, B. Sassolas, D. Stoppa-Lyonnet, L. Thomas, P. Vabres, L. Venat, E. Wierzbicka, and H. Zattara, The contribution of large genomic deletions at the CDKN2A locus to the burden of familial melanoma. Br J Cancer, 2008. 99(2): p. 364–70.

30. Cancer Genome Atlas, N., Genomic Classification of Cutaneous Melanoma. Cell, 2015. 161(7): p. 1681–96.

31. Jonsson, G., C. Dahl, J. Staaf, T. Sandberg, P.O. Bendahl, M. Ringner, P. Guldberg, and A. Borg, Genomic profiling of malignant melanoma using tiling-resolution arrayCGH. Oncogene, 2007. 26(32): p. 4738–48.

32. Peris, K., G. Keller, S. Chimenti, A. Amantea, H. Kerl, and H. Hofler, Microsatellite instability and loss of heterozygosity in melanoma. J Invest Dermatol, 1995. 105(4): p. 625–8.

33. Ohta, M., D. Berd, M. Shimizu, H. Nagai, M.G. Cotticelli, M. Mastrangelo, J.A. Shields, C.L. Shields, C.M. Croce, and K. Huebner, Deletion mapping of chromosome region 9p21-p22 surrounding the CDKN2 locus in melanoma. Int J Cancer, 1996. 65(6): p. 762–7.

34. Kumar, R., J. Smeds, B. Lundh Rozell, and K. Hemminki, Loss of heterozygosity at chromosome 9p21 (INK4-p14ARF locus): homozygous deletions and mutations in the p16 and p14ARF genes in sporadic primary melanomas. Melanoma Res, 1999. 9(2): p. 138–47.

35. Palmieri, G., A. Cossu, P.A. Ascierto, G. Botti, M. Strazzullo, A. Lissia, M. Colombino, M. Casula, C. Floris, F. Tanda, M. Pirastu, G. Castello, and G. Melanoma Cooperative, Definition of the role of chromosome 9p21 in sporadic melanoma through genetic analysis of primary tumours and their metastases. The Melanoma Cooperative Group. Br J Cancer, 2000. 83(12): p. 1707–14.

36. Holland, E.A., S.C. Beaton, B.G. Edwards, R.F. Kefford, and G.J. Mann, Loss of heterozygosity and homozygous deletions on 9p21-22 in melanoma. Oncogene, 1994. 9(5): p. 1361–5.

37. Puig, S., A. Ruiz, C. Lazaro, T. Castel, M. Lynch, J. Palou, A. Vilalta, J. Weissenbach, J.M. Mascaro, and X. Estivill, Chromosome 9p deletions in cutaneous malignant melanoma tumors: the minimal deleted region involves markers outside the p16 (CDKN2) gene. Am J Hum Genet, 1995. 57(2): p. 395–402.

38. Hannon, G.J. and D. Beach, p15INK4B is a potential effector of TGF-beta-induced cell cycle arrest. Nature, 1994. 371(6494): p. 257–61.

39. Kamb, A., N.A. Gruis, J. Weaver-Feldhaus, Q. Liu, K. Harshman, S.V. Tavtigian, E. Stockert, R.S. Day, 3rd, B.E. Johnson, and M.H. Skolnick, A cell cycle regulator potentially involved in genesis of many tumor types. Science, 1994. 264(5157): p. 436–40.

40. McNeal, A.S., K. Liu, V. Nakhate, C.A. Natale, E.K. Duperret, B.C. Capell, T. Dentchev, S.L. Berger, M. Herlyn, J.T. Seykora, and T.W. Ridky, CDKN2B Loss Promotes Progression from Benign Melanocytic Nevus to Melanoma. Cancer Discov, 2015. 5(10): p. 1072–85.

41. Patel, S., C.J. Wilkinson, and E.V. Sviderskaya, Loss of Both CDKN2A and CDKN2B Allows for Centrosome Overduplication in Melanoma. J Invest Dermatol, 2020. 140(9): p. 1837–1846 e1.

42. Platz, A., J. Hansson, E. Mansson-Brahme, B. Lagerlof, S. Linder, E. Lundqvist, P. Sevigny, M. Inganas, and U. Ringborg, Screening of germline mutations in the CDKN2A and CDKN2B genes in Swedish families with hereditary cutaneous melanoma. J Natl Cancer Inst, 1997. 89(10): p. 697–702.

43. Laud, K., C. Marian, M.F. Avril, M. Barrois, A. Chompret, A.M. Goldstein, M.A. Tucker, P.A. Clark, G. Peters, V. Chaudru, F. Demenais, A. Spatz, M.W. Smith, G.M. Lenoir, B. Bressac-de Paillerets, and G. French Hereditary Melanoma Study, Comprehensive analysis of CDKN2A (p16INK4A/p14ARF) and CDKN2B genes in 53 melanoma index cases considered to be at heightened risk of melanoma. J Med Genet, 2006. 43(1): p. 39–47.

44. Landi, M.T., A.M. Goldstein, S. Tsang, D. Munroe, W. Modi, M. Ter-Minassian, R. Steighner, M. Dean, N. Metheny, B. Staats, R. Agatep, D. Hogg, and D. Calista, Genetic susceptibility in familial melanoma from northeastern Italy. J Med Genet, 2004. 41(7): p. 557–66.

45. Casula, M., P.A. Ascierto, A. Cossu, M.C. Sini, S. Tore, M. Colombino, M.P. Satta, A. Manca, C. Rozzo, S.M. Satriano, G. Castello, A. Lissia, F. Tanda, and G. Palmieri, Mutation analysis of candidate genes in melanoma-prone families: evidence of different pathogenetic mechanisms at chromosome 9P21. Melanoma Res, 2003. 13(6): p. 571–9.

46. Yepes, S., M.A. Tucker, H. Koka, Y. Xiao, T. Zhang, K. Jones, A. Vogt, L. Burdette, W. Luo, B. Zhu, A. Hutchinson, M. Yeager, B. Hicks, K.M. Brown, N.D. Freedman, S.J. Chanock, A.M. Goldstein, and X.R. Yang, Integrated Analysis of Coexpression and Exome Sequencing to Prioritize Susceptibility Genes for Familial Cutaneous Melanoma. J Invest Dermatol, 2022. 142(9): p. 2464–2475 e5.

47. Goldstein, A.M., Y. Xiao, J. Sampson, B. Zhu, M. Rotunno, H. Bennett, Y. Wen, K. Jones, A. Vogt, L. Burdette, W. Luo, B. Zhu, M. Yeager, B. Hicks, J. Han, I. De Vivo, S. Koutros, G. Andreotti, L. Beane-Freeman, M. Purdue, N.D. Freedman, S.J. Chanock, L. A. Tucker, and X.R. Yang, Rare germline variants in known melanoma susceptibility genes in familial melanoma. Hum Mol Genet, 2017. 26(24): p. 4886–4895.

48. Artomov, M., A.J. Stratigos, I. Kim, R. Kumar, M. Lauss, B.Y. Reddy, B. Miao, C. Daniela Robles-Espinoza, A. Sankar, C.N. Njauw, K. Shannon, E.S. Gragoudas, A. Marie Lane, V. Iyer, J.A. Newton-Bishop, D. Timothy Bishop, E.A. Holland, G.J. Mann, T. Singh, M.J. Daly, and H. Tsao, Rare Variant, Gene-Based Association Study of Hereditary Melanoma Using Whole-Exome Sequencing. J Natl Cancer Inst, 2017. 109(12).

49. Ivarsdottir, E.V., J. Gudmundsson, V. Tragante, G. Sveinbjornsson, S. Kristmundsdottir, S.N. Stacey, G.H. Halldorsson, M.I. Magnusson, A. Oddsson, G.B. Walters, A. Sigurdsson, S. Saevarsdottir, D. Beyter, G. Thorleifsson, B.V. Halldorsson, P. Melsted, H. Stefansson, I. Jonsdottir, E. Sorensen, O.B. Pedersen, C. Erikstrup, M. Bogsted, M. Pohl, A. Roder, H.V. Stroomberg, I. Gogenur, J. Hillingso, S.E. Bojesen, U. Lassen, E. Hogdall, H. Ullum, S. Brunak, S.R. Ostrowski, D.G. Consortium, I.E. Sonderby, O. Frei, S. Djurovic, A. Havdahl, P. Moller, M. Dominguez-Valentin, J. Haavik, O.A. Andreassen, E. Hovig, B.A. Agnarsson, R. Hilmarsson, O.T. Johannsson, T. Valdimarsson, S. Jonsson, P.H. Moller, J.H. Olafsson, B. Sigurgeirsson, J.G. Jonasson, G. Tryggvason, H. Holm, P. Sulem, T. Rafnar, D.F. Gudbjartsson, and K. Stefansson, Gene-based burden tests of rare germline variants identify six cancer susceptibility genes. Nat Genet, 2024. 56(11): p. 2422–2433.

50. Harland, M., E.A. Holland, P. Ghiorzo, M. Mantelli, G. Bianchi-Scarra, A.M. Goldstein, K.A. Tucker, B.A. Ponder, G.J. Mann, D.T. Bishop, and J. Newton Bishop, Mutation screening of the CDKN2A promoter in melanoma families. Genes Chromosomes Cancer, 2000. 28(1): p. 45–57.

51. Tsubari, M., E. Tiihonen, and M. Laiho, Cloning and characterization of p10, an alternatively spliced form of p15 cyclin-dependent kinase inhibitor. Cancer Res, 1997. 57(14): p. 2966–73.

52. Simon, M., G. Koster, M. Ludwig, R. Mahlberg, S. Rho, M. Watzka, and J. Schramm, Alternative splicing of the p15 cdk inhibitor in glioblastoma multiforme. Acta Neuropathol, 2001. 102(2): p. 167–74.

53. Perez de Castro, I., M. Benet, M. Jimenez, S. Alzabin, M. Malumbres, and A. Pellicer, Mouse p10, an alternative spliced form of p15INK4b, inhibits cell cycle progression and malignant transformation. Cancer Res, 2005. 65(8): p. 3249–56.

54. Cheng, J., G. Novati, J. Pan, C. Bycroft, A. Zemgulyte, T. Applebaum, A. Pritzel, L.H. Wong, M. Zielinski, T. Sargeant, R.G. Schneider, A.W. Senior, J. Jumper, D. Hassabis, P. Kohli, and Z. Avsec, Accurate proteome-wide missense variant effect prediction with AlphaMissense. Science, 2023. 381(6664): p. eadg7492.

55. Zhang, T., J. Choi, M.A. Kovacs, J. Shi, M. Xu, N.C.S. Program, C. Melanoma Meta-Analysis, A.M. Goldstein, A.J. Trower, D.T. Bishop, M.M. Iles, D.L. Duffy, S. MacGregor, L.T. Amundadottir, M.H. Law, S.K. Loftus, W.J. Pavan, and K.M. Brown, Cell-type-specific eQTL of primary melanocytes facilitates identification of melanoma susceptibility genes. Genome Res, 2018. 28(11): p. 1621–1635.

56. Tiffen, J., S.J. Gallagher, F. Filipp, D. Gunatilake, A.A. Emran, C. Cullinane, K. Dutton-Register, L. Aoude, N. Hayward, A. Chatterjee, E.J. Rodger, M.R. Eccles, and P. Hersey, EZH2 Cooperates with DNA Methylation to Downregulate Key Tumor Suppressors and IFN Gene Signatures in Melanoma. J Invest Dermatol, 2020. 140(12): p. 2442–2454 e5.

57. Avsec, Z., N. Latysheva, J. Cheng, G. Novati, K.R. Taylor, T. Ward, C. Bycroft, L. Nicolaisen, E. Arvaniti, J. Pan, R. Thomas, V. Dutordoir, M. Perino, S. De, A. Karollus, A. Gayoso, T. Sargeant, A. Mottram, L.H. Wong, P. Drotar, A. Kosiorek, A. Senior, R. Tanburn, T. Applebaum, S. Basu, D. Hassabis, and P. Kohli, Advancing regulatory variant effect prediction with AlphaGenome. Nature, 2026. 649(8099): p. 1206–1218.

58. Coetzee, S.G., G.A. Coetzee, and D.J. Hazelett, motifbreakR: an R/Bioconductor package for predicting variant effects at transcription factor binding sites. Bioinformatics, 2015. 31(23): p. 3847–9.

59. Coetzee, S.G. and D.J. Hazelett, MotifbreakR v2: extended capability and database integration. ArXiv, 2024.

60. Hashimoto, H., D. Wang, J.R. Horton, X. Zhang, V.G. Corces, and X. Cheng, Structural Basis for the Versatile and Methylation-Dependent Binding of CTCF to DNA. Mol Cell, 2017. 66(5): p. 711–720 e3.

61. Rowlands, C.F., S. Allen, J. Balmana, S.M. Domchek, D.G. Evans, H. Hanson, N. Hoogerbrugge, P.A. James, K.L. Nathanson, M. Robson, M. Tischkowitz, W.D. Foulkes, and C. Turnbull, Population-based germline breast cancer gene association studies and meta-analysis to inform wider mainstream testing. Ann Oncol, 2024. 35(10): p. 892–901.

62. Goldstein, A.M., J. Kim, J.S. Haley, J. Li, D.T. Smelser, H.S. Rao, J.N. Hatton, X.R. Yang, K. Brown, M.A. Tucker, E.G. Berry, A.J. Waters, D.J. Adams, W.M. Linehan, J.M. Kirkwood, P.A. Kanetsky, D.J. Carey, D.R. Stewart, and M.R. Sargen, Familial Melanoma Genes: Prevalence and Cancer Risks Among Genomically Ascertained Individuals. 2026: p. in press.

63. Rao, S.S., M.H. Huntley, N.C. Durand, E.K. Stamenova, I.D. Bochkov, J.T. Robinson, A.L. Sanborn, I. Machol, A.D. Omer, E.S. Lander, and E.L. Aiden, A 3D map of the human genome at kilobase resolution reveals principles of chromatin looping. Cell, 2014. 159(7): p. 1665–80.

64. Thakur, R., M. Xu, H. Sowards, J. Yon, L. Jessop, T. Myers, T. Zhang, R. Chari, E. Long, T. Rehling, R. Hennessey, K. Funderburk, J. Yin, M.J. Machiela, M.E. Johnson, A.D. Wells, A. Chesi, S.F.A. Grant, M.M. Iles, M.T. Landi, M.H. Law, C. Melanoma Meta-Analysis, J. Choi, and K.M. Brown, Mapping chromatin interactions at melanoma susceptibility loci uncovers distant cis-regulatory gene targets. Am J Hum Genet, 2025. 112(7): p. 1625–1648.

65. Kamb, A., D. Shattuck-Eidens, R. Eeles, Q. Liu, N.A. Gruis, W. Ding, C. Hussey, T. Tran, Y. Miki, J. Weaver-Feldhaus, and, et al., Analysis of the p16 gene (CDKN2) as a candidate for the chromosome 9p melanoma susceptibility locus. Nat Genet, 1994. 8(1): p. 23–6.

66. Demenais, F., H. Mohamdi, V. Chaudru, A.M. Goldstein, J.A. Newton Bishop, D.T. Bishop, P.A. Kanetsky, N.K. Hayward, E. Gillanders, D.E. Elder, M.F. Avril, E. Azizi, P. van Belle, W. Bergman, G. Bianchi-Scarra, B. Bressac-de Paillerets, D. Calista, C. Carrera, J. Hansson, M. Harland, D. Hogg, V. Hoiom, E.A. Holland, C. Ingvar, M.T. Landi, J.M. Lang, R.M. Mackie, G.J. Mann, M.E. Ming, C.J. Njauw, H. Olsson, J. Palmer, L. Pastorino, S. Puig, J. Randerson-Moor, M. Stark, H. Tsao, M.A. Tucker, P. van der Velden, X.R. Yang, N. Gruis, and C. Melanoma Genetics, Association of MC1R variants and host phenotypes with melanoma risk in CDKN2A mutation carriers: a GenoMEL study. J Natl Cancer Inst, 2010. 102(20): p. 1568–83.

67. Hnisz, D., A.S. Weintraub, D.S. Day, A.L. Valton, R.O. Bak, C.H. Li, J. Goldmann, B.R. Lajoie, Z.P. Fan, A.A. Sigova, J. Reddy, D. Borges-Rivera, T.I. Lee, R. Jaenisch, M.H. Porteus, J. Dekker, and R.A. Young, Activation of proto-oncogenes by disruption of chromosome neighborhoods. Science, 2016. 351(6280): p. 1454–1458.

68. Rahme, G.J., N.M. Javed, K.L. Puorro, S. Xin, V. Hovestadt, S.E. Johnstone, and B.E. Bernstein, Modeling epigenetic lesions that cause gliomas. Cell, 2023. 186(17): p. 3674–3685 e14.

69. Yang, X.R., M. Rotunno, Y. Xiao, C. Ingvar, H. Helgadottir, L. Pastorino, R. van Doorn, H. Bennett, C. Graham, J.N. Sampson, M. Malasky, A. Vogt, B. Zhu, G. Bianchi-Scarra, W. Bruno, P. Queirolo, G. Fornarini, J. Hansson, R. Tuominen, L. Burdett, B. Hicks, A. Hutchinson, K. Jones, M. Yeager, S.J. Chanock, M.T. Landi, V. Hoiom, H. Olsson, N. Gruis, P. Ghiorzo, M.A. Tucker, and A.M. Goldstein, Multiple rare variants in high-risk pancreatic cancer-related genes may increase risk for pancreatic cancer in a subset of patients with and without germline CDKN2A mutations. Hum Genet, 2016. 135(11): p. 1241–1249.

70. Pathak, A., A. Pemov, M.L. McMaster, R. Dewan, S. Ravichandran, E. Pak, A. Dutra, H.J. Lee, A. Vogt, X. Zhang, M. Yeager, S. Anderson, M. Kirby, N.D.C.G.R. Laboratory, N.D.C.S.W. Group, N. Caporaso, M.H. Greene, L.R. Goldin, and D.R. Stewart, Juvenile myelomonocytic leukemia due to a germline CBL Y371C mutation: 35-year follow-up of a large family. Hum Genet, 2015. 134(7): p. 775–87.

71. Wang, K., M. Li, and H. Hakonarson, ANNOVAR: functional annotation of genetic variants from high-throughput sequencing data. Nucleic Acids Res, 2010. 38(16): p. e164.

72. Poplin, R., P.C. Chang, D. Alexander, S. Schwartz, T. Colthurst, A. Ku, D. Newburger, J. Dijamco, N. Nguyen, P.T. Afshar, S.S. Gross, L. Dorfman, C.Y. McLean, and M.A. DePristo, A universal SNP and small-indel variant caller using deep neural networks. Nat Biotechnol, 2018. 36(10): p. 983–987.

73. Poplin, R., V. Ruano-Rubio, M.A. DePristo, T.J. Fennell, M.O. Carneiro, G.A. Van der Auwera, D.E. Kling, L.D. Gauthier, A. Levy-Moonshine, D. Roazen, K. Shakir, J. Thibault, S. Chandran, C. Whelan, M. Lek, S. Gabriel, M.J. Daly, B. Neale, D.G. MacArthur, and E. Banks, Scaling accurate genetic variant discovery to tens of thousands of samples. bioRxiv, 2018: p. 201178.

74. Martin, M., M. Patterson, S. Garg, S. O Fischer, N. Pisanti, G.W. Klau, A. Schöenhuth, and T. Marschall, WhatsHap: fast and accurate read-based phasing. bioRxiv, 2016: p. 085050.

75. Jiang, T., S. Cao, Y. Liu, Z. Zhang, B. Liu, R. Luo, G. Wang, and Y. Wang, cuteFC: regenotyping structural variants through an accurate and efficient force-calling method. Genome Biol, 2025. 26(1): p. 166.

76. Jiang, T., Y. Liu, Y. Jiang, J. Li, Y. Gao, Z. Cui, Y. Liu, B. Liu, and Y. Wang, Long-read-based human genomic structural variation detection with cuteSV. Genome Biol, 2020. 21(1): p. 189.

77. Xu, M., L. Mehl, T. Zhang, R. Thakur, H. Sowards, T. Myers, L. Jessop, A. Chesi, M.E. Johnson, A.D. Wells, H.T. Michael, P. Bunda, K. Jones, H. Higson, R.C. Hennessey, A. Jermusyk, M.A. Kovacs, M.T. Landi, M.M. Iles, A.M. Goldstein, C. Melanoma Meta-Analysis, J. Choi, S.J. Chanock, S.F.A. Grant, R. Chari, G. Merlino, M.H. Law, and K.M. Brown, A UVB-responsive common variant at chromosome band 7p21.1 confers tanning response and melanoma risk via regulation of the aryl hydrocarbon receptor, AHR. Am J Hum Genet, 2021. 108(9): p. 1611–1630.

78. Dobin, A., C.A. Davis, F. Schlesinger, J. Drenkow, C. Zaleski, S. Jha, P. Batut, M. Chaisson, and T.R. Gingeras, STAR: ultrafast universal RNA-seq aligner. Bioinformatics, 2013. 29(1): p. 15–21.

79. Khanna, A., D.E. Larson, S.N. Srivatsan, M. Mosior, T.E. Abbott, S. Kiwala, T.J. Ley, E.J. Duncavage, M.J. Walter, J.R. Walker, O.L. Griffith, M. Griffith, and C.A. Miller, Bam-readcount - rapid generation of basepair-resolution sequence metrics. ArXiv, 2021.

80. Poulos, R.C., J.A.I. Thoms, Y.F. Guan, A. Unnikrishnan, J.E. Pimanda, and J.W.H. Wong, Functional Mutations Form at CTCF-Cohesin Binding Sites in Melanoma Due to Uneven Nucleotide Excision Repair across the Motif. Cell Rep, 2016. 17(11): p. 2865–2872.

81. Liu, Q., J.A.I. Thoms, A.C. Nunez, Y. Huang, K. Knezevic, D. Packham, R.C. Poulos, R. Williams, D. Beck, N.J. Hawkins, R.L. Ward, J.W.H. Wong, L.B. Hesson, M.A. Sloane, and J.E. Pimanda, Disruption of a −35 kb Enhancer Impairs CTCF Binding and MLH1 Expression in Colorectal Cells. Clin Cancer Res, 2018. 24(18): p. 4602–4611.

82. Vasimuddin, M., S. Misra, H. Li, and S. Aluru, Efficient Architecture-Aware Acceleration of BWA-MEM for Multicore Systems. 2019 Ieee 33rd International Parallel and Distributed Processing Symposium (Ipdps 2019), 2019: p. 314–324.

83. Li, H., B. Handsaker, A. Wysoker, T. Fennell, J. Ruan, N. Homer, G. Marth, G. Abecasis, R. Durbin, and S. Genome Project Data Processing, The Sequence Alignment/Map format and SAMtools. Bioinformatics, 2009. 25(16): p. 2078–9.

84. Ramirez, F., D.P. Ryan, B. Gruning, V. Bhardwaj, F. Kilpert, A.S. Richter, S. Heyne, F. Dundar, and T. Manke, deepTools2: a next generation web server for deep-sequencing data analysis. Nucleic Acids Res, 2016. 44(W1): p. W160–5.

85. Katainen, R., K. Dave, E. Pitkanen, K. Palin, T. Kivioja, N. Valimaki, A.E. Gylfe, H. Ristolainen, U.A. Hanninen, T. Cajuso, J. Kondelin, T. Tanskanen, J.P. Mecklin, H. Jarvinen, L. Renkonen-Sinisalo, A. Lepisto, E. Kaasinen, O. Kilpivaara, S. Tuupanen, M. Enge, J. Taipale, and L.A. Aaltonen, CTCF/cohesin-binding sites are frequently mutated in cancer. Nat Genet, 2015. 47(7): p. 818–21.

86. Belaghzal, H., J. Dekker, and J.H. Gibcus, Hi-C 2.0: An optimized Hi-C procedure for high-resolution genome-wide mapping of chromosome conformation. Methods, 2017. 123: p. 56–65.

87. Langmead, B., C. Wilks, V. Antonescu, and R. Charles, Scaling read aligners to hundreds of threads on general-purpose processors. Bioinformatics, 2019. 35(3): p. 421–432.

88. Langmead, B. and S.L. Salzberg, Fast gapped-read alignment with Bowtie 2. Nat Methods, 2012. 9(4): p. 357–9.

89. Langmead, B., C. Trapnell, M. Pop, and S.L. Salzberg, Ultrafast and memory-efficient alignment of short DNA sequences to the human genome. Genome Biol, 2009. 10(3): p. R25.

90. Servant, N., N. Varoquaux, B.R. Lajoie, E. Viara, C.J. Chen, J.P. Vert, E. Heard, J. Dekker, and E. Barillot, HiC-Pro: an optimized and flexible pipeline for Hi-C data processing. Genome Biol, 2015. 16: p. 259.

91. Ernst, J. and M. Kellis, Chromatin-state discovery and genome annotation with ChromHMM. Nat Protoc, 2017. 12(12): p. 2478–2492.

92. Zeng, H., A. Jorapur, A.H. Shain, U.E. Lang, R. Torres, Y. Zhang, A.S. McNeal, T. Botton, J. Lin, M. Donne, I.N. Bastian, R. Yu, J.P. North, L. Pincus, B.S. Ruben, N.M. Joseph, I. Yeh, B.C. Bastian, and R.L. Judson, Bi-allelic Loss of CDKN2A Initiates Melanoma Invasion via BRN2 Activation. Cancer Cell, 2018. 34(1): p. 56–68 e9.

93. Bycroft, C., C. Freeman, D. Petkova, G. Band, L.T. Elliott, K. Sharp, A. Motyer, D. Vukcevic, O. Delaneau, J. O’Connell, A. Cortes, S. Welsh, A. Young, M. Effingham, G. McVean, S. Leslie, N. Allen, P. Donnelly, and J. Marchini, The UK Biobank resource with deep phenotyping and genomic data. Nature, 2018. 562(7726): p. 203–209.

94. Conroy, M.C., B. Lacey, J. Besevic, W. Omiyale, Q. Feng, M. Effingham, J. Sellers, S. Sheard, M. Pancholi, G. Gregory, J. Busby, R. Collins, and N.E. Allen, UK Biobank: a globally important resource for cancer research. Br J Cancer, 2023. 128(4): p. 519–527.

95. Robinson, J.T., H. Thorvaldsdottir, W. Winckler, M. Guttman, E.S. Lander, G. Getz, and J.P. Mesirov, Integrative genomics viewer. Nat Biotechnol, 2011. 29(1): p. 24–6.

